# Autism heterogeneity related to preterm birth: multi-ancestry results from the SPARK sample

**DOI:** 10.1101/2025.06.05.25328988

**Authors:** Charikleia Chatzigeorgiou, Zeynep Asgel, Marina Natividad Avila, Behrang Mahjani, Vahe Khachadourian, Tade Souaiaia, Niamh Mullins, Magdalena Janecka

## Abstract

Autism Spectrum Disorder (ASD) shows significant clinical variability, likely due to a combination of genetic and environmental factors. Preterm birth is a known risk factor for ASD, occurring in approximately 13% of diagnosed individuals. While genetic factors contribute to preterm birth in the general population, the relationship between genetic variation, preterm birth, and ASD heterogeneity remains unclear.

We investigated the genetic factors associated with preterm birth in autistic individuals using data from the SPARK sample. We conducted three ancestry-specific, genome-wide association studies for African/African American, Admixed American, and Non-Finnish European ancestries, followed by a meta-analysis using METAL. Functional mapping and gene-based analyses were performed using FUMA, and genetic correlations were estimated using LDSC and Popcorn. Polygenic risk scores (PRS) were computed with BridgePRS.

Our study identified ancestry-specific genetic loci associated with preterm birth in ASD cases. Although the meta-analysis results were not statistically significant, the estimated SNP heritability was 14%, indicating a meaningful contribution of common genetic variants. Across ancestry groups, preterm birth status was not significantly associated with PRS for any psychiatric or medical conditions analyzed. However, polygenic liability to preterm birth was linked to several congenital anomalies after multiple testing adjustments.

These findings underline the importance of including diverse ancestries in genetic studies of preterm birth in ASD and underscore the potential for utilizing early-life exposure information to understand ASD heterogeneity. Future research should replicate these findings in larger samples and explore rare variants associated with preterm birth to better understand the relationship between gestational duration and clinical and genetic differences in ASD.

## Introduction

Approximately 1% of children worldwide[1], and 1 in 36 children in the US[2], are diagnosed with autism spectrum disorder (ASD). ASD is characterized by differences in social communication and restricted/repetitive patterns of behavio [3], and is often accompanied by multiple co-occurring conditions, including attention-deficit/hyperactivity disorder (ADHD), motor delay and anxiety disorders[4]. Beyond the core diagnostic symptoms, autism is highly heterogeneous, which manifests at multiple levels: its etiology, age of onset, clinical symptoms and their progression, as well as the pattern of co-occurring conditions. However, both causes of autism, and the factors contributing to case heterogeneity remain largely unknown[5] — and outside of research on monogenic forms of autism[6–8], few studies have explored the mapping between distinct etiological factors and clinically-defined subgroups of cases.

While the *causal* role of early-life exposures in autism remains unclear[9], our previous research has shown their utility in identifying subsets of cases with distinct pattern of co-occurring conditions[4]. For example, individuals born preterm were more likely to be diagnosed with difficulty in gaining weight or motor delay (compared to autistic individuals born to term), while those whose mothers reported infection in pregnancy were more likely to suffer from hearing loss[4]. These exposures were also more likely to occur in autistic individuals than their unaffected siblings. Therefore, irrespective of the causal vs. non-causal role of *e.g.,* preterm birth in autism, information on the preterm status can contribute to prediction of autism risk[10] and can be leveraged to study autism heterogeneity.

Given our earlier results demonstrating *clinical* differences among autistic individuals based on their early-life exposures[4], here we focus on one of such exposures – preterm birth – to better understand the mechanisms underlying the link between early exposures and clinical variability of autism. Preterm birth is relatively common [11], and it is defined as the delivery before completing 37 weeks of gestation. Preterm birth is associated with a number of health consequences for the child, spectrum and intensity of which vary, with the risk escalating the earlier the child is delivered[12]. The sequelae of preterm birth extend beyond the immediate newborn phase, underscoring the imperative need for comprehensive care and support to navigate the diverse health consequences associated with early deliveries.

As the duration of pregnancy in the general population is influenced in part by fetal genetic factors[13], we investigated the presence of systematic genetic differences between autistic individuals born term and preterm, and the degree to which those differences can account for the phenotypic heterogeneity reported previously in autism. We used a multi-ancestry SPARK cohort to detect genetic loci differentiating autistic individuals born term and preterm, and then performed their functional analysis, and studied the genetic and phenotypic overlap between preterm birth and other traits. Collectively, such analyses can help shed light on the genetic landscape associated with preterm birth in autism, elucidate genetic factors contributing to case heterogeneity, and unravel the intricate relationships between preterm birth and other phenotypes in autistic individuals.

## Methods

### Sample

Simons Foundation Powering Autism Research for Knowledge (SPARK) is a large autism study, with both online and in-person enrollment. Eligibility criteria include a diagnosis of ASD and residence in the US. Participants are encouraged to enroll with their parents and siblings, irrespective of their ASD status. Upon enrolment, all participants and/or their caregivers complete a series of demographic, cognitive, behavioral and medical questionnaires[14]. DNA is extracted from saliva samples. Genotype data was derived from the Illumina Global Screening Array (GSA v1, v2, GSA_24v2-0_A2).

We used phenotypic and genetic data drawn from SPARK iWES v2 data freeze. Preterm birth was defined as self-or caregiver-reported delivery before 37 weeks of gestation. Individuals with an ASD diagnosis born prematurely were defined as *cases*. Individuals with a diagnosis of ASD born to term were defined as *controls.*

### Genotyping and imputation

#### Pre-imputation quality control

We first removed SNPs located within sex or mitochondrial chromosomes, or with low genotyping rate (<0.9). Subsequently, we removed samples with high genotype missingness (>0.1) and discrepancies between self-reported and genotype-derived sex. We retained SNPs with genotyping rate ≥ 0.98, minor allele frequency (MAF) ≥ 0.01, deviation from Hardy-Weinberg equilibrium (HWE) p ≥ 10^-6^ in cases or p ≥ 10^-10^ in controls. Additionally, we removed related individuals, one from each pair with Identity-By-Descent (PI_HAT)>0.2, retaining the individual with higher call rates within the pair.

#### Imputation and Post-imputation QC

Input data preparation for imputation was performed according to the data preparation guidelines provided by TOPMed Imputation Server. Quality control for TOPMed reference panel was carried out using the toolbox provided by Will Rayner (http://www.well.ox.ac.uk/~wrayner/tools/). Due to diverse ancestry of the SPARK individuals, all genotypes were imputed to a diverse reference panel (TOPMed panel; 194,512 haplotypes; version R2 on GRCh38). Genotype imputation for each chromosome was performed after the standard QC and phasing steps. Post-imputation QC was performed using PLINK v1.9 and vcftools [15] by extracting SNPs with MAF ≥ 0.01, info score ≥ 0.5, genotype missingness ≤ 1%, and Hardy-Weinberg equilibrium p-value ≥ 1 × 10^−^□.

#### Ancestry mapping

We adopted the ancestry mapping approach developed by gnomAD (https://gnomad.broadinstitute.org/news/2023-11-the_mgenetic-ancestry/). The dataset was combined with the HGDP + 1KG subset of gnomAD and restricted to 5,000 ancestry-informative SNPs[16]. Principal Component Analysis (PCA) was performed in the joint dataset and a Random Forest classifier was trained on the first 10 Principal Components of HGDP + 1KG samples to infer ancestry for the SPARK samples, ensuring consistency in the classification process. Only SPARK samples from Admixed American (AMR), African/African American (AFR) and Non-Finnish European (EUR) continental ancestry labels were retained.

#### Genome-wide association analysis (GWAS) and meta-analysis

GWAS of preterm birth was done using the imputed genotype dosage data with PLINK v2.0 and VCF tools. PCA was performed separately for each ancestry group using LD-pruned, genotyped SNPs to generate eigenvectors. We conducted three ancestry-specific GWAS [AFR, AMR, EUR], in each adjusting for the 15 ancestry-specific Principal Components (note: using 10 Principal Components resulted in a highly deflated genomic control factor). We then conducted a meta-analysis of the ancestry-specific GWASs using standard error fixed effect model with genomic control correction implemented in METAL (thereafter referred to as multi-ancestry GWAS)[17]. For the downstream analyses, we utilized full summary statistics without applying LD-based clumping. Since these methods incorporate their own LD correction strategies, pre-clumping was not necessary, enabling us to retain all informative SNPs.

#### Post-GWAS analysis

#### Functional annotation

We conducted annotation, functional mapping, gene-based analysis and gene-set analysis of multi-ancestry GWAS using SNP2GENE function module of FUMA[18]. For gene prioritization we selected all genes and adopted the default settings otherwise (maximum P-value of lead SNPs of 5×10^−6^; r^2^ threshold for independent significant SNPs of 0.6). Subsequently, we used both mapped genes and the significant genes from gene-based analysis to construct a gene expression heatmap via GENE2FUNC function, enabling us to explore the expression of preterm-associated genes across tissues. All analyses were run across ancestries using a multi-ancestry reference panel (1000 Genome Project phase 3[19]). GSEA function (FUMA) was used to explore the biological functions of the prioritized genes and compare them with genes in the GWAS catalog[20] and gene sets in Molecular Signatures Database (MsigDB) v7.1[21]. Overall, there were 20,260 background genes applied to GSEA. Gene sets were reported if they met two criteria: (i) at least two prioritized genes in the gene set; (ii) the adjusted *P*-value of the gene set was□<□0.05.

#### Heritability and genetic correlations of preterm birth and other conditions and traits

To estimate SNP-based heritability of preterm birth in ASD cases and to calculate genetic correlations between preterm birth in our ASD sample and other traits, we used LDSC for analyses involving only European ancestry data (EUR-EUR correlations)[22] and Popcorn for analyses involving Admixed American and African ancestry data in comparison to European ancestry (AMR/AFR-EUR correlations)[23]. While LDSC enabled us to compute genetic correlations across ancestry-matched European individuals in our and external datasets, Popcorn is tailored for cross-population genetic correlation analysis, implementing weighted likelihood function that accounts for the inflation of Z-scores due to LD to provide robust estimates even when LD patterns differ between populations. The traits included in the analyses were: epilepsy[24], ADHD[25], anorexia nervosa[26], gastrointestinal disease[27], bipolar disorder[28], schizophrenia (SCZ) [29], musculoskeletal system disease[27], immune system disease[27] and panic disorder[30]. All summary statistics were sourced from online databases and subsequently converted to the hg38 genome assembly.

As input for the LDSC analysis, we used summary statistics from European-ancestry GWAS of preterm birth in the SPARK sample. Prior to the analyses, we filtered SNPs to those found in individuals from the 1000 Genomes Project and converted the files to LDSC format (implemented in munge_sumstats.py). We focused our analysis on well-imputed SNPs by filtering to those found in the HapMap3 reference panel[31]. The pre-computed LD scores[32], with major histocompatibility complex (MHC) region excluded, were used for the analyses for individuals of European ancestry.

Genotype data for the EUR, AMR, and AFR populations in Popcorn analyses were sourced from the 1000 Genomes Project Phase 3[19]. Prior to analysis, standard quality control procedures were applied using PLINK, with SNPs with MAF > 0.01, non-autosomal chromosomes and the MHC region removed. Analyses were performed for both AMR and AFR ancestry individuals, as well as the full meta-analyzed sample. To address the issue of extreme results, the top 5% of betas, which represented significant outliers in the GWAS results, were identified and removed. The distributions of beta coefficients before and after this filtering process are provided in the supplemental materials **(Fig. S1)**, highlighting the changes and the rationale for the exclusion of these outliers.

#### Polygenic risk score (PRS) analyses

To test the genetic overlap between preterm birth and other traits in our sample, we (1) tested the association between polygenic risk score (PRS) for preterm birth and other phenotypes available for individuals in the SPARK cohort. Additionally, we (2) calculated PRS for additional phenotypes (both psychiatric and non-psychiatric) in the SPARK sample and tested their association with the preterm birth *status* reported for the SPARK participants.

To compute the PRS across the traits and ancestries, we used BridgePRS [33], an advanced Bayesian PRS method tailored to enhance the transferability of PRS across diverse ancestral backgrounds. Each ancestry was analyzed separately to account for ancestry-specific genetic architectures. Clumping was performed with parameters set to 1000 kb and an LD r2 threshold of 0.01 to ensure the independence of SNPs. The LD reference panel used was based on 1000 Genomes variants with a minor allele frequency (MAF) greater than 1% from the corresponding population. Additionally, the analysis included the first 15 principal components as covariates.

#### Associations between polygenic liability to preterm birth and other phenotypes

We analyzed the association between polygenic liability (PRS) to preterm birth and an array of medical conditions captured through the medical screening module in SPARK. Preterm birth PRS was estimated for all autistic individuals in our analyses, irrespective of preterm status, using external preterm base data (EUR) (N= 84,689) [34] to avoid inflating the associations between PRS and other phenotypes. Medical history module data in SPARK is based on self-or parental report of clinical diagnosis and is divided into several domains. A positive reply for each domain is followed by more specific and detailed questions regarding the conditions within that domain. We tested the association between preterm PRS and 68 medical conditions in the domains of (a) birth or pregnancy complications, (b) attention or behavioral disorders, (c) developmental disabilities, (d) growth conditions, (e) neurological conditions, (f) vision or hearing conditions, (g) psychiatric disorders, and (h) sleep, feeding, eating or toileting problems. Each of the medical conditions within these domains served as an outcome in a separate model. PRS score was standardized prior to analyses. Additional model covariates included the individual’s sex and year of birth. Given the pre-imputation QC, the individuals were unlikely to be genetically related, however, to address the potential non-genetic correlation between individuals from the same family, the standard errors were estimated using clustered sandwich estimator, implemented in the clusterSEs package (v2.6.2).

#### Association between preterm birth status and polygenic liability to other traits

Polygenic risk scores (PRSs) for additional traits were calculated in BridgePRS using relevant summary statistics from EUR, AFR and AMR ancestries, as available. **Table S1** provides details on the datasets used for each trait and ancestry.

## Results

Genotype data for 647,280 variants from 69,758 individuals with genotype data (56.8% males) were available from the SPARK project [35]. We excluded 32,884 without a confirmed ASD diagnosis. After genotyping quality control, 400,266 variants and data from 31,947 individuals were retained, including 3,308 autistic individuals born prematurely and 28,639 born at term. After imputation, there were a total of 8,855,081 variants with MAF>0.01 and imputation quality score (INFO)>0.5.

### Genetic differences between ASD cases born term and preterm

Classification into three ancestry groups provided the best fit for the data, and individuals who were highly admixed were excluded from the analyses, leaving 2,888 preterm cases in the analysis. We conducted three ancestry-specific GWASs [African/African American (AFR; n_preterm_=196, n_term_=1,722), Admixed American (AMR; n_preterm_=396, n_term_=3,955), Non-Finnish European (EUR; n_preterm_=2,296, n_term_=19,173)] adjusting for the 15 principal components.

In ancestry-specific GWASs, only the AFR GWAS had SNPs reaching genome-wide level of significance (p<5.0×10^-08^; rs117965482 (chr 12), p=3.04×10^-08^ and rs78395263 (chr 12), p=3.89×10^-08^; **Fig. S2**). The top associated variant from in AMR GWAS was rs4713424 (chr 6, p=1.30×10^-07^ (**Fig. S3**) and in EUR GWAS rs114444 (chr 16, p=1.45×10^-07^; **Fig. S4**). The direction of the effects of those loci on preterm birth differed across ancestries (*e.g.,* rs78395263 (effect allele T) had an allele frequency of 3.6% in EUR and 1.6% in AFR, with EUR - OR=0.998, P=0.978 and AFR - OR=5.40, P=3.89×10×□□.; rs117965482 (effect allele C) had an allele frequency of 0.4% in EUR and 0.5% in AFR, with EUR - OR=0.992, P=0.920 and AFR - OR=5.50, P=3.04×10×□□; **Fig. S5**). The QQ plots for each GWAS can be found in **Fig. S6.**

The differential direction of the effect sizes rendered the variants identified in ancestry-specific GWAS non-significant in the meta-analysis. The top associated variant in the meta-analysis was rs2019941, effect allele T (OR=0.71 (0.62-0.81), P=4.52 × 10^-07^) on chromosome 8 (**Fig. 1**).

**Figure 1.**
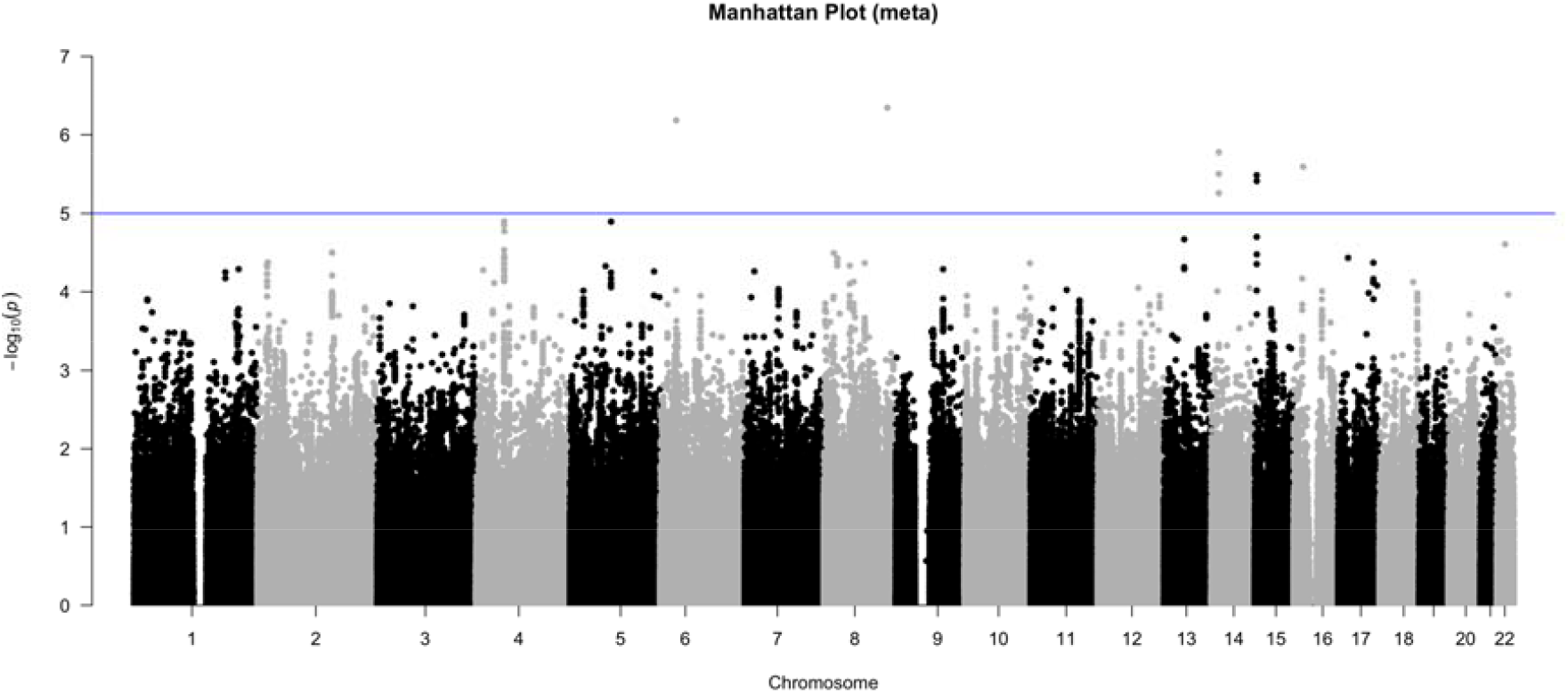
Manhattan plot of the results from the logistic regression of the metanalysis of preterm (n=3,308) vs. non-preterm (n=28,639) ASD cases, after adjustment for the first 15 PCs. Genomic regions contain SNPs that exceed 5 × 10^-05^ above the blue line. No locus in the meta-analyzed GWAS was genome-wide significant.

### Functional annotation of the preterm-associated loci in ASD

In FUMA analyses of the multi-ancestry meta, we identified four genomic risk loci associated with preterm birth in ASD cases at P < 5.0×10^-6^, located on chromosomes 8, 14, 15, and 16 (**Figure S7).** These loci include the SNPs rs2019941 (chr8:129126122, p = 4.52 × 10×□), rs118182680 (chr14:35462315, p = 1.67 × 10×□), rs58406701 (chr15:28114815, p = 3.24 × 10?□), and rs114444 (chr16:19609590, p = 2.54 × 10×□). After LD pruning, four independently significant SNPs were retained. Additionally, 59 candidate SNPs were identified via positional mapping, 12 of which were correlated with previously reported loci through LD, suggesting a shared genetic architecture with related traits. However, no direct trait annotations for these loci were available in the GWAS Catalog, suggesting that their functional significance in preterm birth and ASD has not been established. Most candidate SNPs were intronic, supporting a potential role for non-coding regulatory mechanisms in preterm birth susceptibility (**Fig. S8**). Functional annotation based on positional mapping identified three genes related to preterm birth in ASD (*SRP54, C16orf62*, and *OCA2*).

Tissue enrichment analysis of these three genes using MAGMA revealed nominally significant enrichment in nerve tibial (p=0.043) and artery aorta (p = 0.047) tissues among 53 specific tissue types (**Figure S9A**). When grouped into 30 general tissue types, the results revealed significant enrichment in the nerve tissue (p = 0.035), as well as reproductive tissues such as the uterus (p = 0.038) and ovary (p = 0.045) (**Figure S9B**).

To further investigate tissue-specific gene expression, we analyzed differential gene expression patterns using GTEx v8 across both 30 general and 54 specific tissue types. The analysis did not reveal any tissues exhibiting statistically significant differential expression after multiple testing correction. The highest level of gene expression enrichment was in the testis. A heatmap of average expression levels for *SRP54, C16orf62*, and *OCA2* across various tissues highlighted distinct tissue-specific expression patterns. SRP54 exhibited high levels of tissue-specific expression, whereas *OCA2*, on the other hand, displayed relatively uniform expression levels across tissues. *C16orf62* showed a range of expression levels across tissue types (**Figure S10**).

### Genetic overlap between preterm birth and other phenotypes

SNP-based heritability of preterm birth in the full, multi-ancestry sample calculated with LDSC was significant and higher than reported in the general population (14.24%, SE=0.023, Mean Chi^2^: 1.04), cf. 2.5% reported in the general population [34]; see **Fig. S11** for heritability estimates across ancestries). SNP-based heritability of preterm birth in the EUR ancestry calculated with LDSC was statistically significant (35.4%, SE=0.027, Mean Chi^2^: 1.03).

The genetic correlation between preterm birth in our ASD sample and an external sample drawn from the general population was non-significant, both in the full, multi-ancestry sample (rg=-0.099, p= 0.720) and the European ancestry (rg=0.105, p=0.548) subset of the SPARK individuals.

### Genetic correlations

Genetic correlations between preterm birth in our ASD sample and other traits estimated for the meta-analyzed and EUR ancestry individuals in the LDSC analysis were all non-significant except for a negative correlation with SCZ (EUR) (**Table S2**; **Figure 2**). However, in the meta-analyzed sample analyzed with Popcorn, four out of twelve genetic correlations between preterm birth in ASD cases and other traits were significant, all of them negative. The strongest correlations included those between preterm birth and SCZ (cross-ancestry) (P = 0.001), SCZ (EUR) (P = 0.002), bipolar disorder (P = 0.007) and gastrointestinal disease (P = 0.025) (**Figure 2**; **Table S5**; ancestry-specific genetic correlations are presented in **Tables S3-S4**).

**Fig. 2.**
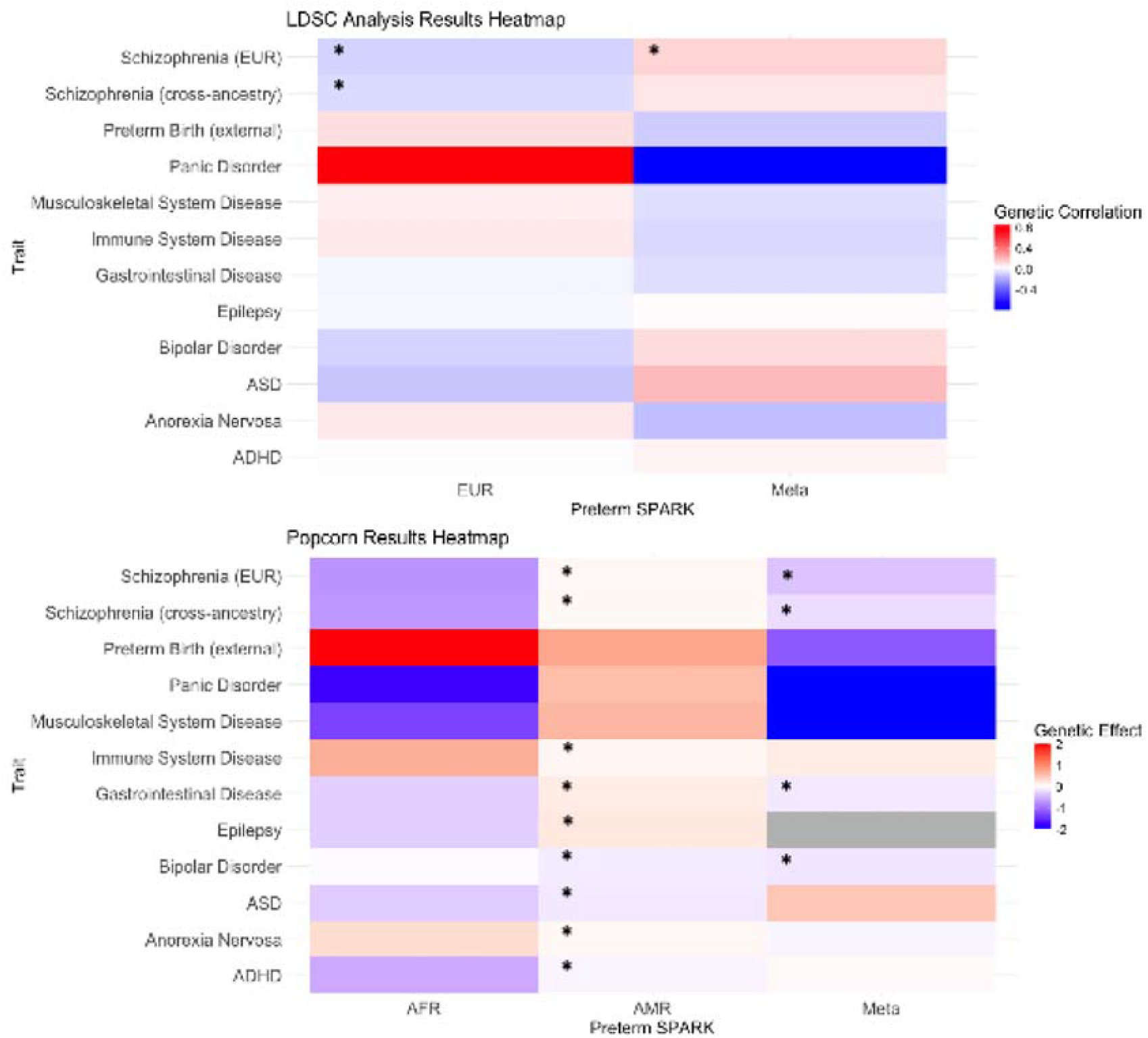
Heatmap illustrating the genetic correlations and effects between traits as estimated using LDSC and Popcorn analyses. Each cell represents the correlation and effect between two traits, with the color gradient ranging from blue (low correlation) to red (high correlation). Significant genetic correlations and effects are indicated by a star. To improve the visual clarity of the heatmap, the color scale was adjusted by clipping genetic effects outside the range of −2 to 2.

### Polygenic risk score analyses

#### Polygenic liability to preterm birth and reported medical conditions

Polygenic liability to preterm birth (PRS) computed using BridgePRS was nominally significantly associated with 4 medical conditions reported in the SPARK medical screening, including birth defects and mood or psychiatric conditions, (**Fig. S11, Table S6**). After adjusting for multiple testing, 3 conditions, including missing kidney (OR = 2.36, 95%CI = 2.24-2.48), esophageal atresia (OR = 0.69, 95%CI = 0.67-0.70), missing uterus (OR = 0.41, 95%CI = 0.39-0.42), remained statistically significant, although the direction of the effects differed across these associations.

#### Preterm birth and genetic liability to medical conditions

Reported preterm birth status was not significantly associated with genetic liability to any of the additional traits we analyzed in any of the ancestry-specific analyses (**Tables S7-S8**; **Fig. S13**).

## Discussion

We examined the genetic underpinnings of preterm birth in autistic individuals, building on our previous work demonstrating clinical heterogeneity among them in association with different early-life exposures[4]. Our results demonstrate that the differences in comorbid conditions in individuals diagnosed with ASD born preterm vs. term co-occur with genetic differences between those groups — highlighting the potential of integrating genetic and early-life information to understand autism heterogeneity.

Our findings revealed ancestry-specific SNPs significantly associated with preterm birth (rs78395263 and rs117965482 for AFR), however, none of these associations remained significant in the trans-ancestry meta-analysis. This suggests that genetic effects on preterm birth might vary between populations – as suggested previously [36, 37] – and that combining data from diverse ancestries could weaken these effects [38, 39]. It is possible that these genetic factors operate through environmental influences in an ancestry-specific manner – as reported previously for other traits [40]. While such environment-mediated effects could be largely maternal in origin, we were not able to separate distinct maternal and fetal effects on the risk of preterm birth[34, 41].

The gene regions mapped to significant SNPs through positional mapping - *SRP54, OCA2*, and *C16orf62* (also called VPS35L) did not reach genome-wide significance in gene-based tests. These genes have been previously linked to congenital neutropenia [42] (*SPR54*), pigmentation and vision [43] (*OCA2*), and Ritscher-Schinzel syndrome [44, 45] - a neurodevelopmental condition associated with a host of comorbid conditions, including alterations in the skeletal, gastrointestinal, immune and genitourinary systems [46]

Leveraging the genome-wide signals in the SPARK and general population samples, we observed that the genetic correlation of preterm birth in individuals with ASD and the general population [34] was not significant, indicating that the genetic factors contributing to preterm birth might differ in those with autism. While the GWAS signal in autistic individuals might be enriched for pleiotropic loci – affecting both autism and preterm birth risk – compared to the signal in the general population, the power to uncover a genetic correlation between those two was limited by the low SNP heritability, warranting further replication. The low SNP heritability estimates of preterm birth reported in our and previous studies stand in contrast to twin- and family-based estimates – 30-40% [47] - likely indicating the effects of rare variants not captured by common SNPs. As these rare variants are often missed by standard SNP arrays, heritability estimates based on common SNPs can underestimate the true genetic variance [40, 48, 49], and limit the statistical power in studies like ours. Alternatively, the differences in heritability estimates might be caused by its overestimation in family studies due to shared environmental effects or non-additive genetic variation [48].

Further analyses revealed genetic overlap between preterm birth and other traits and conditions in autistic individuals. These correlations differed across the ancestry groups, potentially due to different genetic variants associated with preterm birth [36, 37] or the distinct LD patterns in AMR and AFR individuals that may affect the ability to detect genetic correlations [50]. Environmental factors or population-specific interactions could also further separate the genetic risks for preterm birth across ancestries. In the meta-analyzed sample, we found significant genetic correlations with schizophrenia (EUR) using LDSC, as well as schizophrenia (both EUR and cross-ancestry), gastrointestinal disease, and bipolar disorder using Popcorn. The differences in results between LDSC and Popcorn could be attributed to how each method estimates genetic association. LDSC is influenced by factors such as sample size, LD structure, and the genetic architecture of the trait. In contrast, Popcorn accounts for ancestry-specific effects and the extent of shared genetic variation across populations. Additionally, lower heritability in some ancestries might reduce the power to detect genetic correlations with LDSC.

Finally, PRS analyses offered us an independent opportunity to evaluate the genetic overlap between preterm birth in ASD and other phenotypes. Using BridgePRS [33] to calculate polygenic risk across the traits enabled us to implement these analyses across all ancestries in our sample. Our results highlight that polygenic liability to preterm birth is associated with co-occurring conditions, especially birth defects. However, some of these associations were in the opposite direction than expected – we observed lower risk of some birth defects in individuals with higher preterm PRS, despite higher frequency of congenital problems in preterm individuals in the general population [51]-indicating caution is warranted when interpreting these associations. Additionally, the lack of association between preterm birth PRS and preterm birth status in the SPARK sample suggests that the genetic factors contributing to preterm birth may differ in individuals with ASD compared to the general population. This aligns with our finding that the genetic correlation between preterm birth in ASD and in the general population was not significant, further supporting the idea that distinct genetic mechanisms may underlie preterm birth in individuals with autism.

In conclusion, we utilized a large and diverse cohort of individuals with an ASD diagnosis, with comprehensive medical and genotype data. We implemented methods suitable for analysis of data from individuals of diverse ancestries throughout our project, allowing us to include the entire sample with relevant information. Our results underline the importance of considering ancestry-specific genetic factors when studying complex traits like preterm birth in ASD. The observed phenotypic and genetic associations suggest the potential of genetic factors associated with early-life exposures to account for clinical heterogeneity in autism. However, while our study supports a proof-of-principle concept – that early-life exposure data can be used to probe genetic differences between autistic individuals, and map those differences onto phenotypic heterogeneity – the power in our study was limited, partly due to the genetic architecture of preterm birth, with substantial contribution of rare variation [52–54]. Future studies should aim to replicate these findings in larger cohorts, accounting for both common and rare variation, including structural variants [54].

## Data Availability

All data produced in the present study are available upon reasonable request to the authors.

## Acknowledgements

We would like to acknowledge the support of the Seaver Foundation Fellowship to Dr. Chatzigeorgiou and the Beatrice and Samuel A. Seaver Foundation to Dr. Mahjani and Ms. Natividad Avila. All other authors report no biomedical financial interests or potential conflicts of interest.

## Code Availability

All analytical code is available at https://github.com/zasgel/PretermHeterogeneity

## References

1. Zeidan, J., et al., Global prevalence of autism: A systematic review update. Autism Res, 2022. 15(5): p. 778–790.

2. Maenner, M.J., et al., Prevalence and Characteristics of Autism Spectrum Disorder Among Children Aged 8 Years - Autism and Developmental Disabilities Monitoring Network, 11 Sites, United States, 2020. MMWR Surveill Summ, 2023. 72(2): p. 1–14.

3. Sandin, S., et al., The Heritability of Autism Spectrum Disorder. JAMA, 2017. 318(12): p. 1182–1184.

4. Khachadourian, V., et al., Comorbidities in autism spectrum disorder and their etiologies. Transl Psychiatry, 2023. 13(1): p. 71.

5. Andreassen, O.A., et al., New insights from the last decade of research in psychiatric genetics: discoveries, challenges and clinical implications. World Psychiatry, 2023. 22(1): p. 4–24.

6. Grove, J., et al., Identification of common genetic risk variants for autism spectrum disorder. Nat Genet, 2019. 51(3): p. 431–444.

7. Rylaarsdam, L. and A. Guemez-Gamboa, Genetic Causes and Modifiers of Autism Spectrum Disorder. Front Cell Neurosci, 2019. 13: p. 385.

8. Jenner, L., et al., Heterogeneity of Autism Characteristics in Genetic Syndromes: Key Considerations for Assessment and Support. Curr Dev Disord Rep, 2023. 10(2): p. 132–146.

9. Khachadourian, V., et al., Familial confounding in the associations between maternal health and autism. Nat Med, 2025.

10. McGowan, E.C. and S.J. Sheinkopf, Autism and Preterm Birth: Clarifying Risk and Exploring Mechanisms. Pediatrics, 2021. 148(3).

11. Ohuma, E.O., et al., National, regional, and global estimates of preterm birth in 2020, with trends from 2010: a systematic analysis. Lancet, 2023. 402(10409): p. 1261–1271.

12. Clinic, M. Premature birth. 2024; Available from: https://www.mayoclinic.org/diseases-conditions/premature-birth/symptoms-causes/syc-20376730.

13. Workalemahu, T., et al., Genetic and Environmental Influences on Fetal Growth Vary during Sensitive Periods in Pregnancy. Sci Rep, 2018. 8(1): p. 7274.

14. SFARI | SPARK Phenotypic Measures. 4/1/2025]; Available from: https://www.sfari.org/spark-phenotypic-measures/.

15. Danecek, P., et al., The variant call format and VCFtools. Bioinformatics, 2011. 27(15): p. 2156–8.

16. Purcell, S.M., et al., A polygenic burden of rare disruptive mutations in schizophrenia. Nature, 2014. 506(7487): p. 185–90.

17. Willer, C.J., Y. Li, and G.R. Abecasis, METAL: fast and efficient meta-analysis of genomewide association scans. Bioinformatics, 2010. 26(17): p. 2190–1.

18. Functional Mapping and Annotation of Genome-wide association studies. 4/1/2025]; Available from: https://fuma.ctglab.nl/.

19. Genomes Project, C., et al., A global reference for human genetic variation. Nature, 2015. 526(7571): p. 68–74.

20. MacArthur, J., et al., The new NHGRI-EBI Catalog of published genome-wide association studies (GWAS Catalog). Nucleic Acids Res, 2017. 45(D1): p. D896–D901.

21. Liberzon, A., et al., Molecular signatures database (MSigDB) 3.0. Bioinformatics, 2011. 27(12): p. 1739–40.

22. Bulik-Sullivan, B., et al., An atlas of genetic correlations across human diseases and traits. Nat Genet, 2015. 47(11): p. 1236–41.

23. Brown, B.C., et al., Transethnic genetic-correlation estimates from summary statistics. The American Journal of Human Genetics, 2016. 99(1): p. 76–88.

24. International League Against Epilepsy Consortium on Complex, E., GWAS meta-analysis of over 29,000 people with epilepsy identifies 26 risk loci and subtype-specific genetic architecture. Nat Genet, 2023. 55(9): p. 1471–1482.

25. Demontis, D., et al., Genome-wide analyses of ADHD identify 27 risk loci, refine the genetic architecture and implicate several cognitive domains. Nat Genet, 2023. 55(2): p. 198–208.

26. Watson, H.J., et al., Genome-wide association study identifies eight risk loci and implicates metabo-psychiatric origins for anorexia nervosa. Nat Genet, 2019. 51(8): p. 1207–1214.

27. Donertas, H.M., et al., Common genetic associations between age-related diseases. Nat Aging, 2021. 1(4): p. 400–412.

28. Mullins, N., et al., Genome-wide association study of more than 40,000 bipolar disorder cases provides new insights into the underlying biology. Nat Genet, 2021. 53(6): p. 817–829.

29. Trubetskoy, V., et al., Mapping genomic loci implicates genes and synaptic biology in schizophrenia. Nature, 2022. 604(7906): p. 502–508.

30. Forstner, A.J., et al., Genome-wide association study of panic disorder reveals genetic overlap with neuroticism and depression. Mol Psychiatry, 2021. 26(8): p. 4179–4190.

31. International HapMap, C., et al., Integrating common and rare genetic variation in diverse human populations. Nature, 2010. 467(7311): p. 52–8.

32. Finucane, H.K., et al., Partitioning heritability by functional annotation using genome-wide association summary statistics. Nat Genet, 2015. 47(11): p. 1228–35.

33. Hoggart, C.J., et al., BridgePRS leverages shared genetic effects across ancestries to increase polygenic risk score portability. Nat Genet, 2024. 56(1): p. 180–186.

34. Liu, X., et al., Variants in the fetal genome near pro-inflammatory cytokine genes on 2q13 associate with gestational duration. Nat Commun, 2019. 10(1): p. 3927.

35., S.C.E.a. and S. Consortium, SPARK: A US Cohort of 50,000 Families to Accelerate Autism Research. Neuron, 2018. 97(3): p. 488–493.

36. Modi, B.P., et al., Discovery of rare ancestry-specific variants in the fetal genome that confer risk of preterm premature rupture of membranes (PPROM) and preterm birth. BMC Med Genet, 2018. 19(1): p. 181.

37. Rappoport, N., et al., A genome-wide association study identifies only two ancestry specific variants associated with spontaneous preterm birth. Sci Rep, 2018. 8(1): p. 226.

38. Zhang, J., et al., Similarity and diversity of genetic architecture for complex traits between East Asian and European populations. BMC Genomics, 2023. 24(1): p. 314.

39. Troubat, L., et al., Multi-trait GWAS for diverse ancestries: mapping the knowledge gap. BMC Genomics, 2024. 25(1): p. 375.

40. Yang, J., et al., Concepts, estimation and interpretation of SNP-based heritability. Nat Genet, 2017. 49(9): p. 1304–1310.

41. Svensson, A.C., et al., Maternal effects for preterm birth: a genetic epidemiologic study of 630,000 families. Am J Epidemiol, 2009. 170(11): p. 1365–72.

42. Bellanne-Chantelot, C., et al., Mutations in the SRP54 gene cause severe congenital neutropenia as well as Shwachman-Diamond-like syndrome. Blood, 2018. 132(12): p. 1318–1331.

43. Donnelly, M.P., et al., A global view of the OCA2-HERC2 region and pigmentation. Hum Genet, 2012. 131(5): p. 683–96.

44. Otsuji, S., et al., Clinical diversity and molecular mechanism of VPS35L-associated Ritscher-Schinzel syndrome. J Med Genet, 2023. 60(4): p. 359–367.

45. Healy, M.D., et al., Structure of the endosomal Commander complex linked to Ritscher-Schinzel syndrome. Cell, 2023. 186(10): p. 2219–2237 e29.

46. Ritscher-Schinzel syndrome. [cited 2025; Available from: https://rarediseases.info.nih.gov/diseases/5666/disease.

47. Wadon, M., et al., Recent advances in the genetics of preterm birth. Ann Hum Genet, 2020. 84(3): p. 205–213.

48. Young, A.I., Solving the missing heritability problem. PLoS Genet, 2019. 15(6): p. e1008222.

49. Genin, E., Missing heritability of complex diseases: case solved? Hum Genet, 2020. 139(1): p. 103–113.

50. Li, Y.R. and B.J. Keating, Trans-ethnic genome-wide association studies: advantages and challenges of mapping in diverse populations. Genome Med, 2014. 6(10): p. 91.

51. Purisch, S.E., et al., Preterm birth in pregnancies complicated by major congenital malformations: a population-based study. Am J Obstet Gynecol, 2008. 199(3): p. 287 e1–8.

52. Huusko, J.M., et al., Whole exome sequencing reveals HSPA1L as a genetic risk factor for spontaneous preterm birth. PLoS Genet, 2018. 14(7): p. e1007394.

53. Modi, B.P., et al., Rare mutations and potentially damaging missense variants in genes encoding fibrillar collagens and proteins involved in their production are candidates for risk for preterm premature rupture of membranes. PLoS One, 2017. 12(3): p. e0174356.

54. Wong, H.S., et al., Contribution of de novo and inherited rare CNVs to very preterm birth. J Med Genet, 2020. 57(8): p. 552–557.

