## Supplemental Material for "Autism heterogeneity related to preterm birth: multi-ancestry results from the SPARK sample"

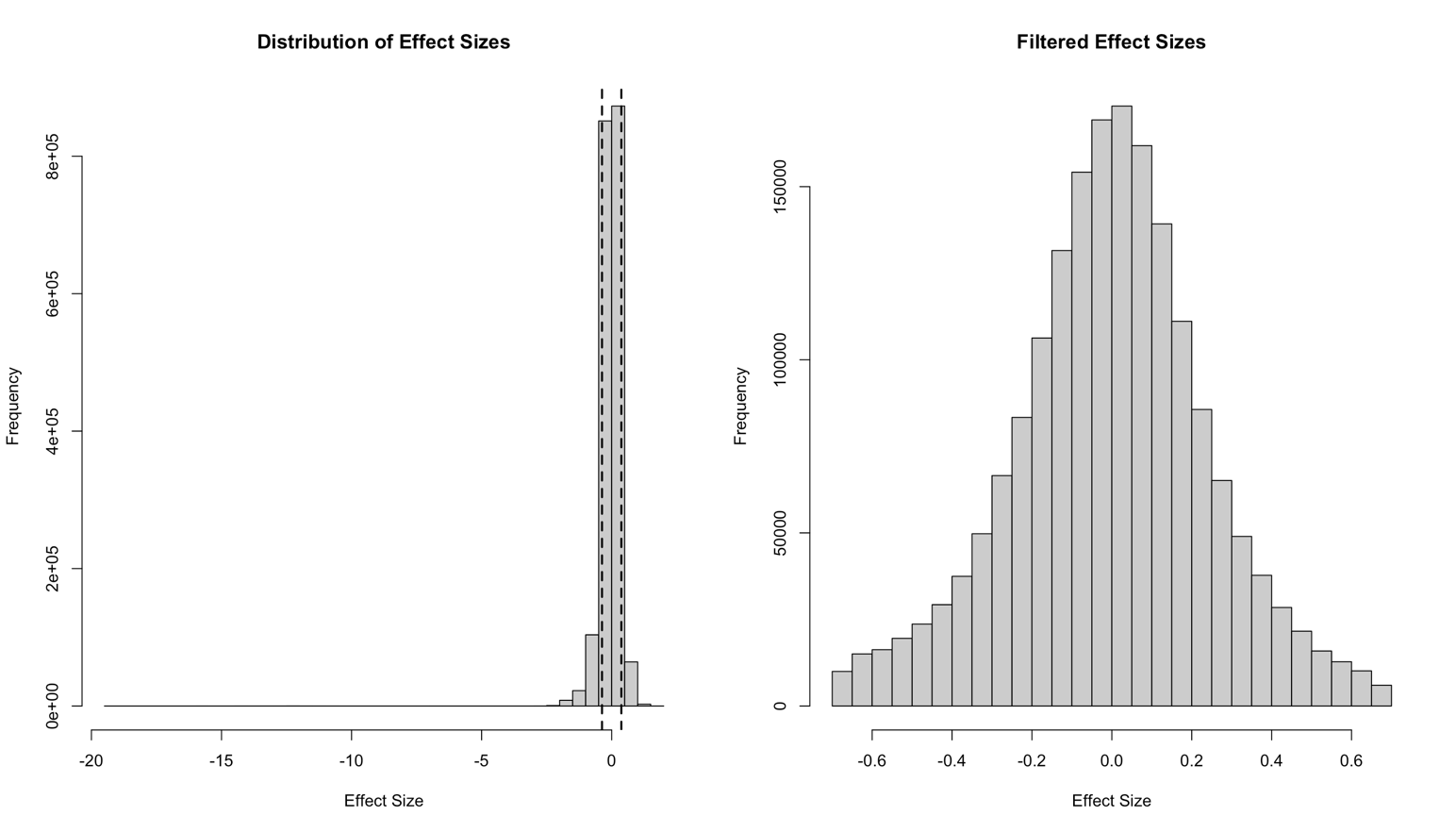

**Fig. S1.** The distribution of effect sizes is shown before filtering (left panel) and after filtering (right panel). In the left panel, the wide range of effect sizes includes extreme outliers. A dashed line indicates the cut-off value of 0.69, corresponding to the top 5% of effect sizes, which were removed during filtering. After filtering (right panel), the distribution exhibits a more symmetrical and centered pattern, highlighting the removal of outliers and the refined focus on relevant effect sizes for downstream analyses.

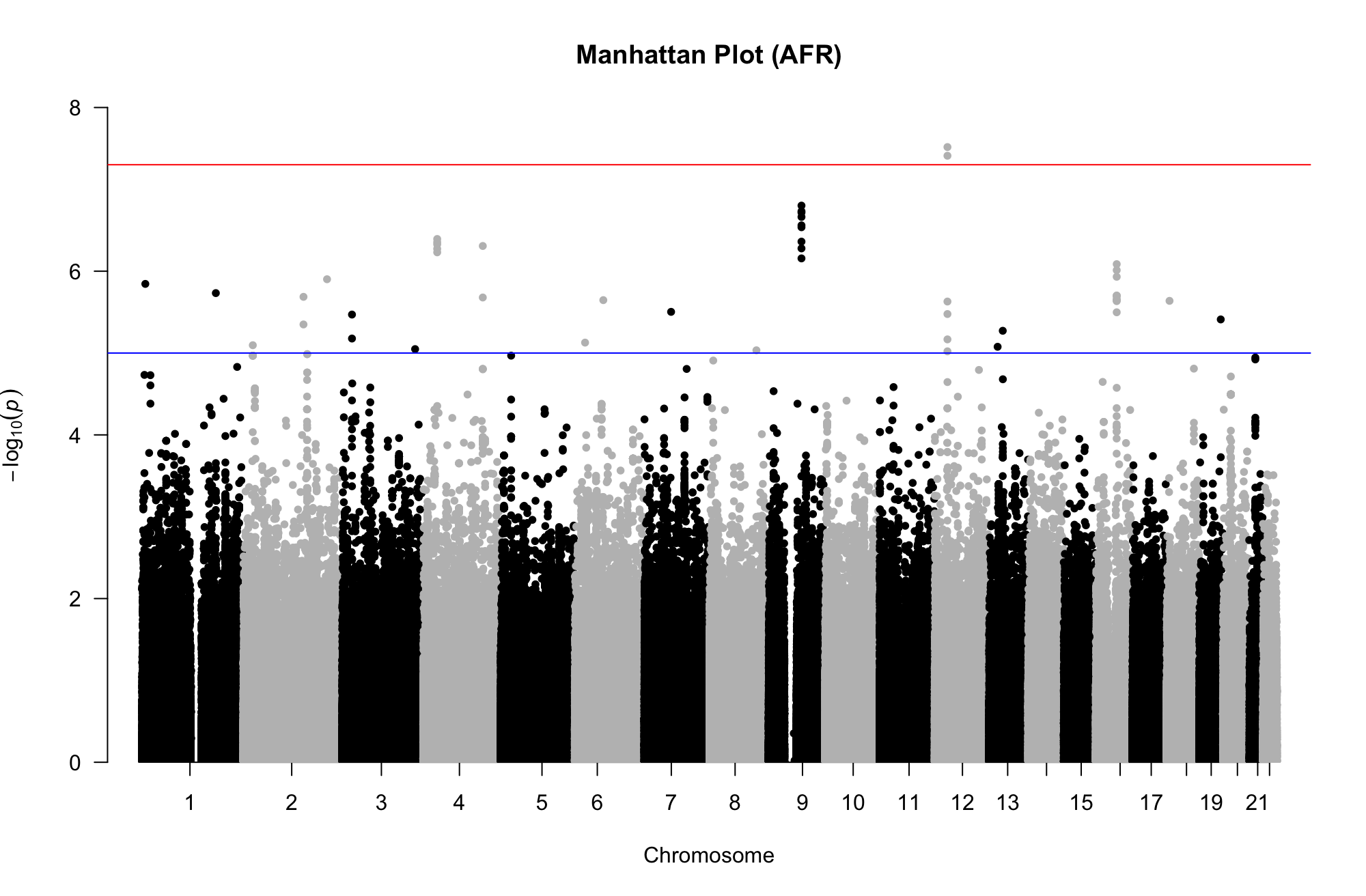
**Figure S2.** Manhattan plot of the results from the logistic regression of preterm (n=196) vs. non-preterm (n=1,722) ASD cases of African/African-American ancestry, after adjustment for the first 15 ancestry-specific PCs. Genomic regions contain SNPs that exceed i) the genome-wide significance threshold of 5 x 10^-08^ are shown above the red line and ii) 5 x 10^-05^ above the blue line.

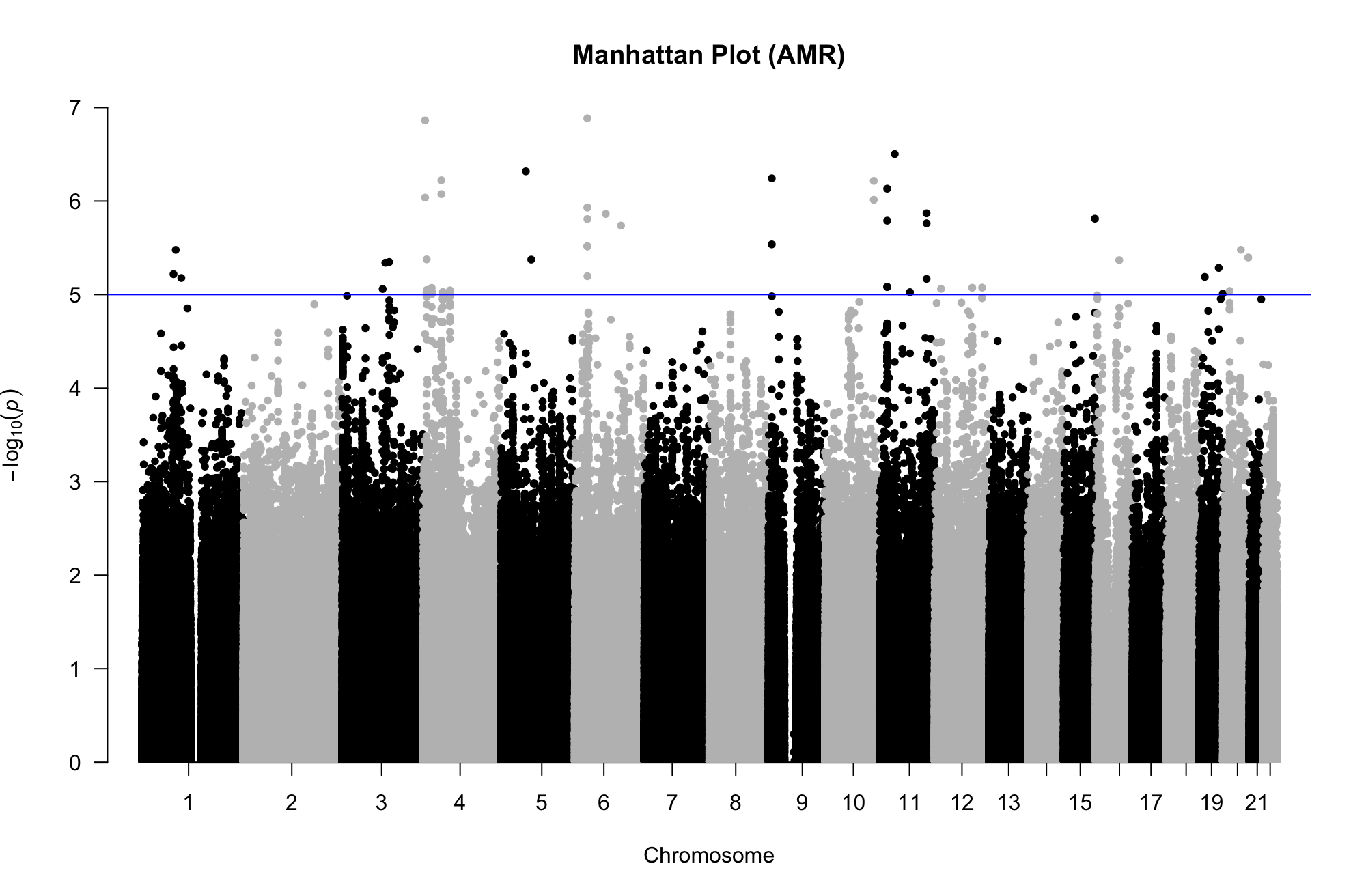

**Figure S3.** Manhattan plot of the results from the logistic regression of preterm (n=396) vs. non-preterm (n=3,955) ASD cases of Admixed American ancestry, after adjustment for the first 15 ancestry-specific PCs. Genomic regions contain SNPs that exceed 5 x 10^-05^ above the blue line. No locus in AMR GWAS was genome-wide significant.

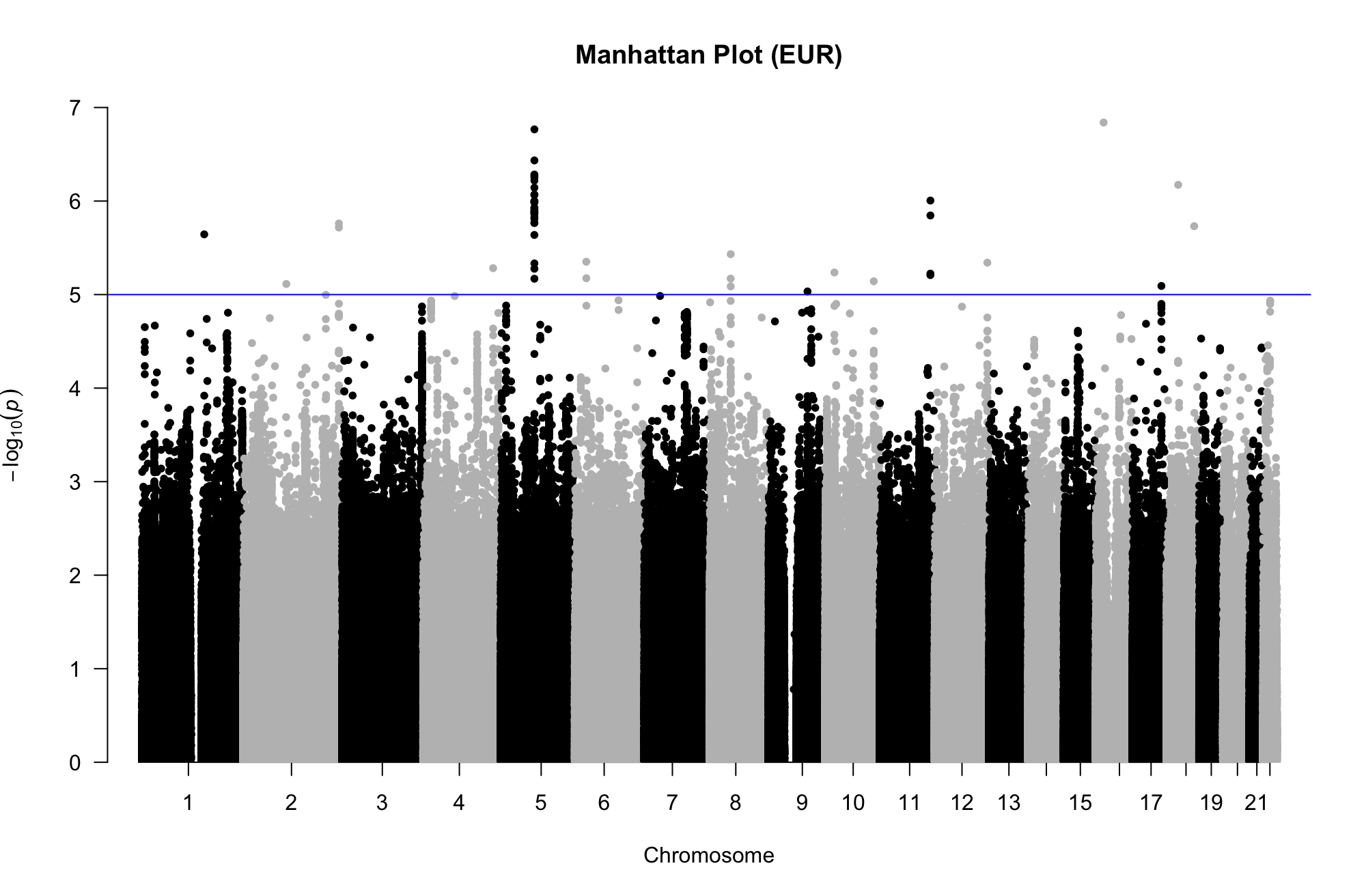

**Figure S4.** Manhattan plot of the results from the logistic regression of preterm (n=2,296) vs. non-preterm (n=19,173) ASD cases of Non-Finish European ancestry after adjustment for the first 15 ancestry-specific PCs. Genomic regions contain SNPs that exceed 5 x 10^-05^ above the blue line. No locus in EUR GWAS was genome-wide significant.

A)
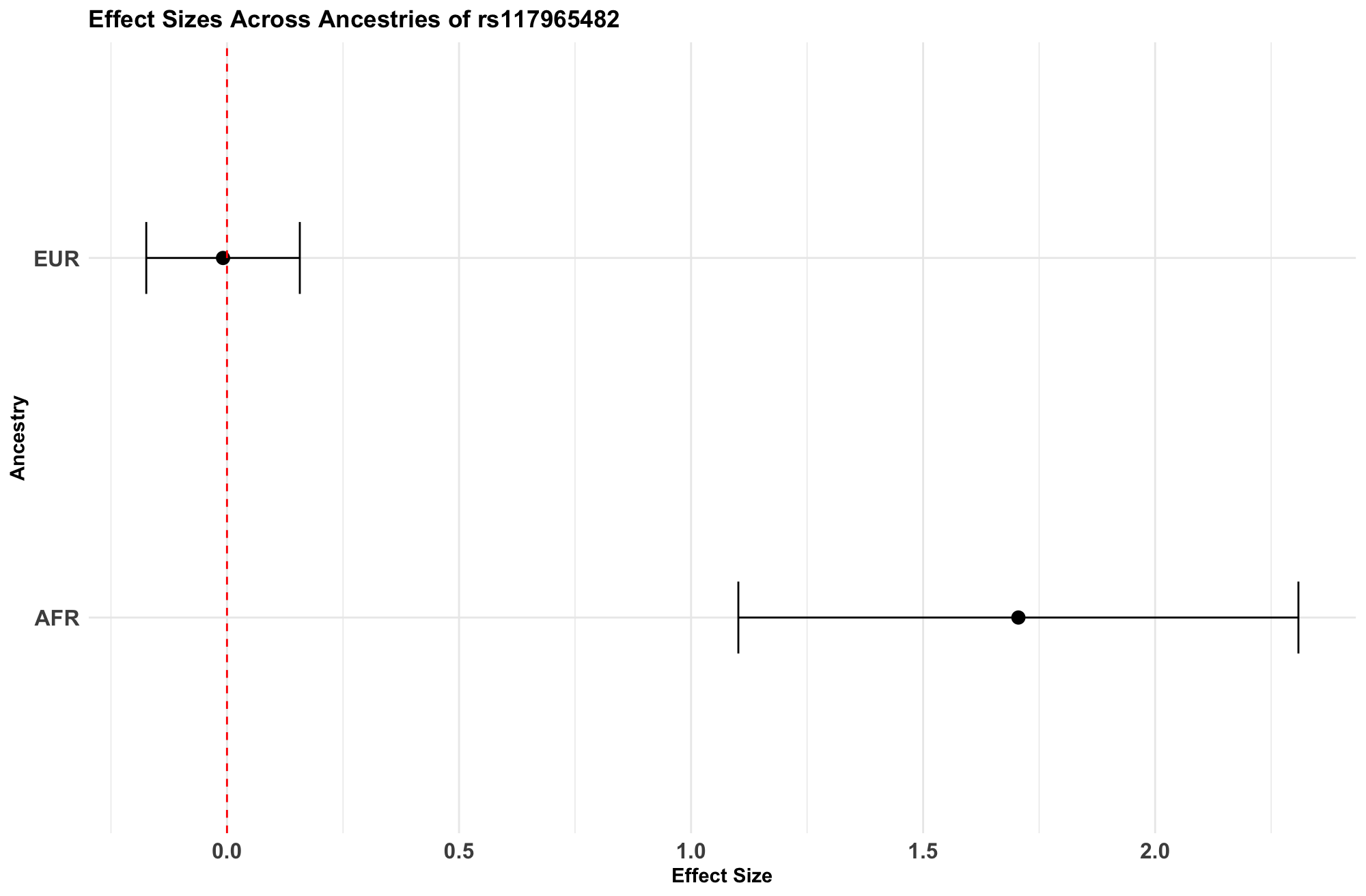

B)
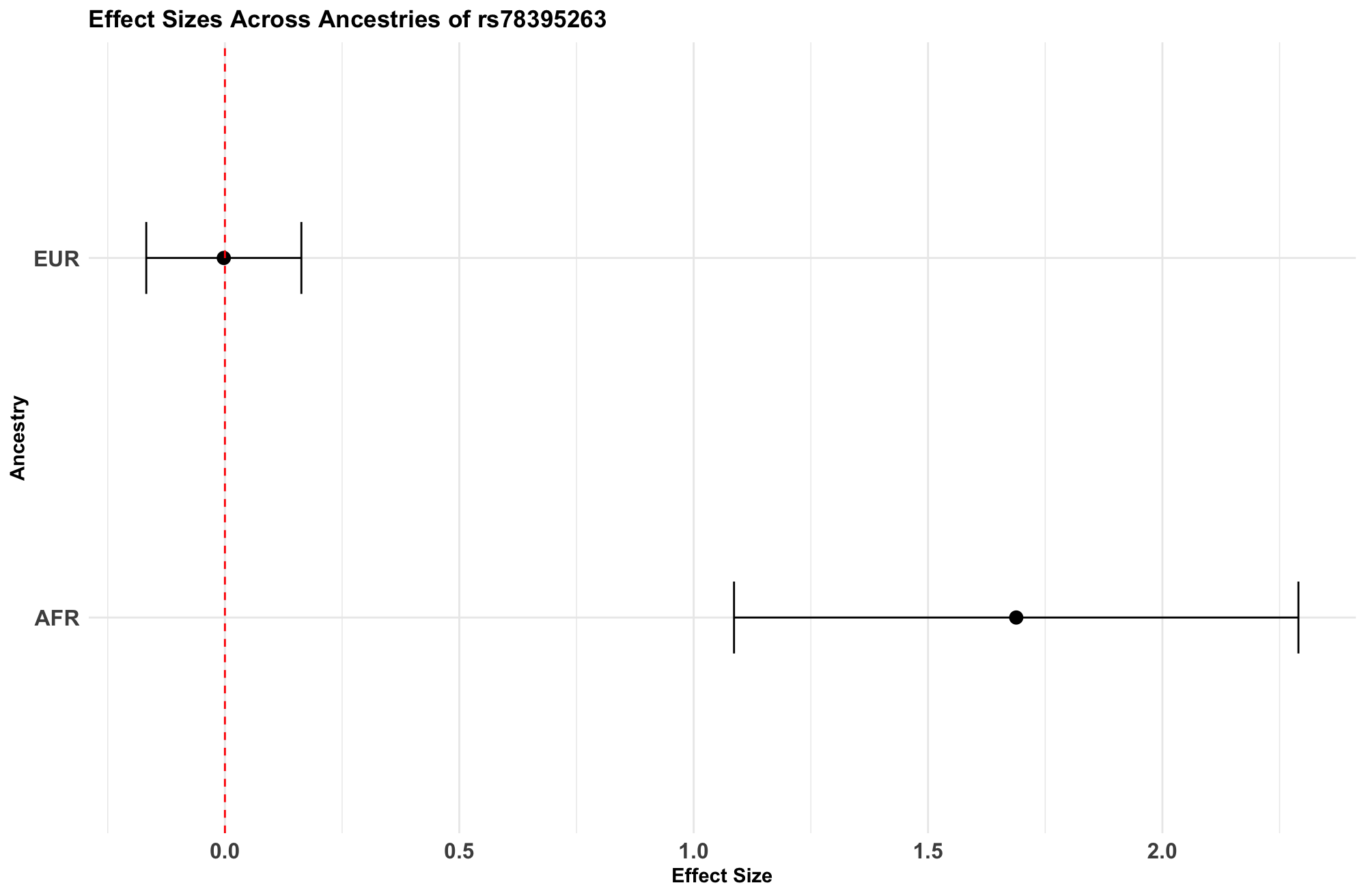

**Fig. S5.** Comparison of Effect Sizes Across EUR and AFR Populations for genome-wide significant SNPs. This figure presents the effect sizes of two genome-wide significant SNPs across European (EUR) and African (AFR) ancestry groups. Panel A shows the effect size distribution for SNP rs117956482 (effect allele C), while Panel B illustrates the effect size for SNP rs78395263 (effect allele T). The x-axis represents the effect size (beta), with error bars indicating confidence intervals. The red dashed line marks the null effect for comparison. The AMR ancestry and meta-analysis did not contain these SNPS.

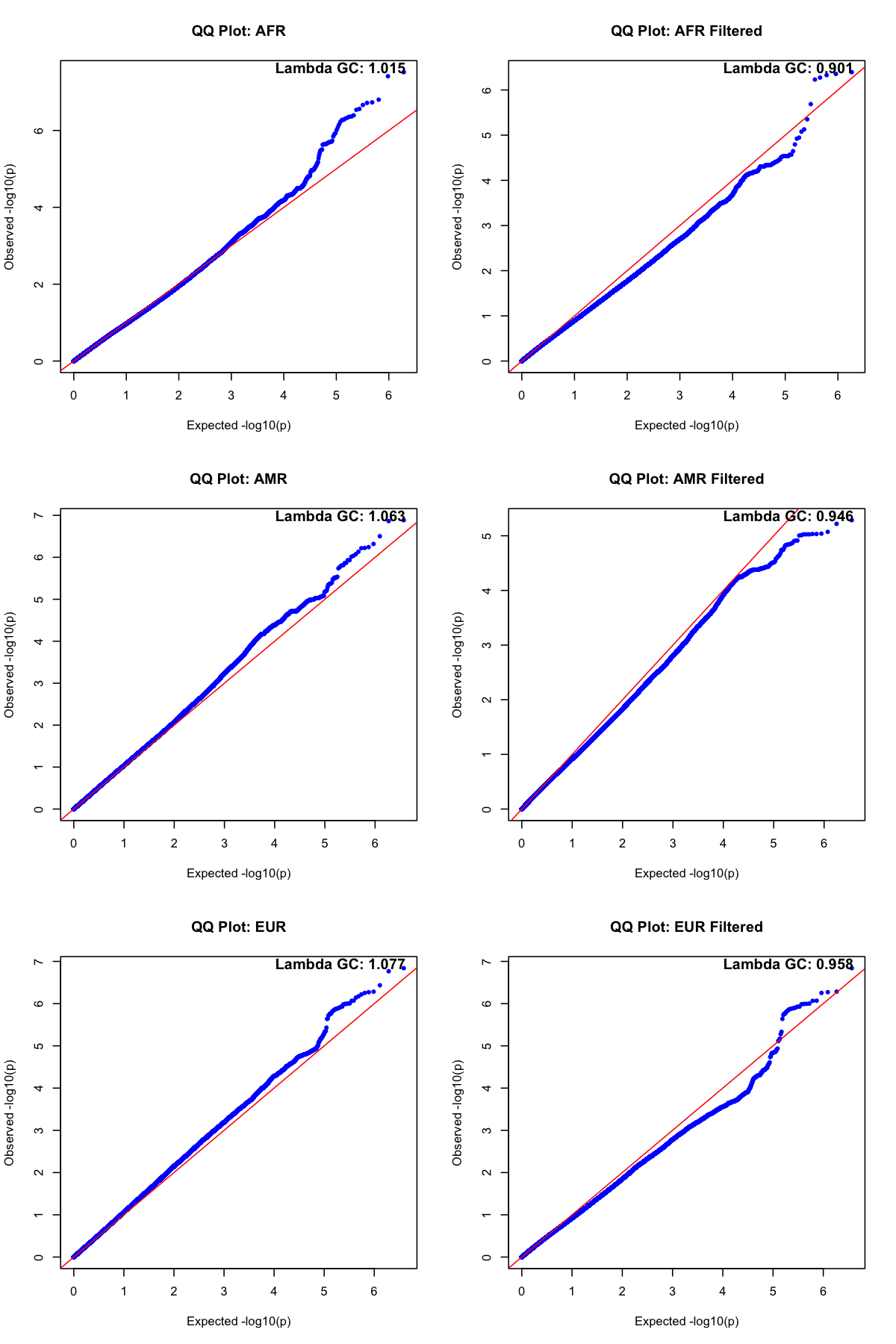

**Fig S6.** QQ plots of ancestry specific GWAS both before and after filtering. The red diagonal line represents the expected distribution, while deviations indicate potential inflation or deflation of test statistics. The genomic control factor (Lambda GC) is reported for each dataset to assess overall inflation.

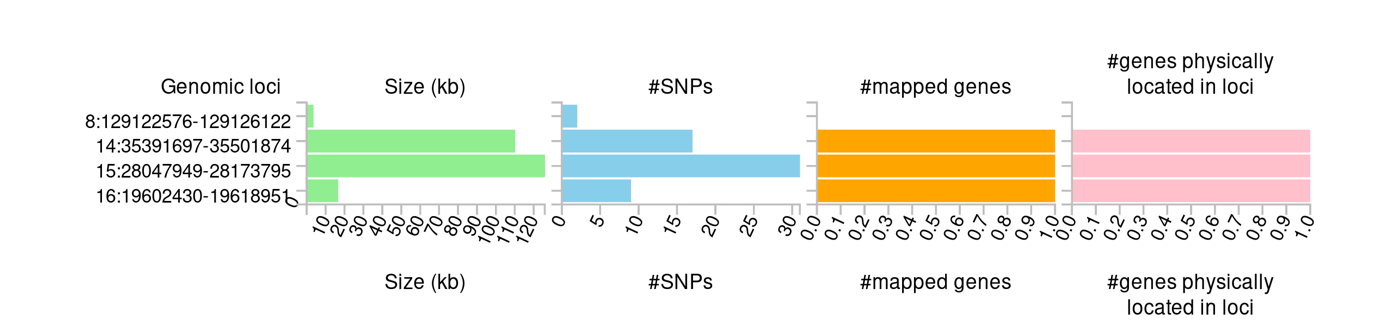

**Fig S7.** Summary of genomic risk loci analysis based on GWAS of ASD preterm individuals. Genomic risk loci are displayed by the ‘chromosome:start position-end position’ on the Y axis. Histograms from left to right depict the size of the genomic locus, number of candidate SNPs in the genomic locus, number of mapped genes by positional mapping and eQTL mapping in the genomic locus, and the number of genes known to be located within the genomic loci, respectively.

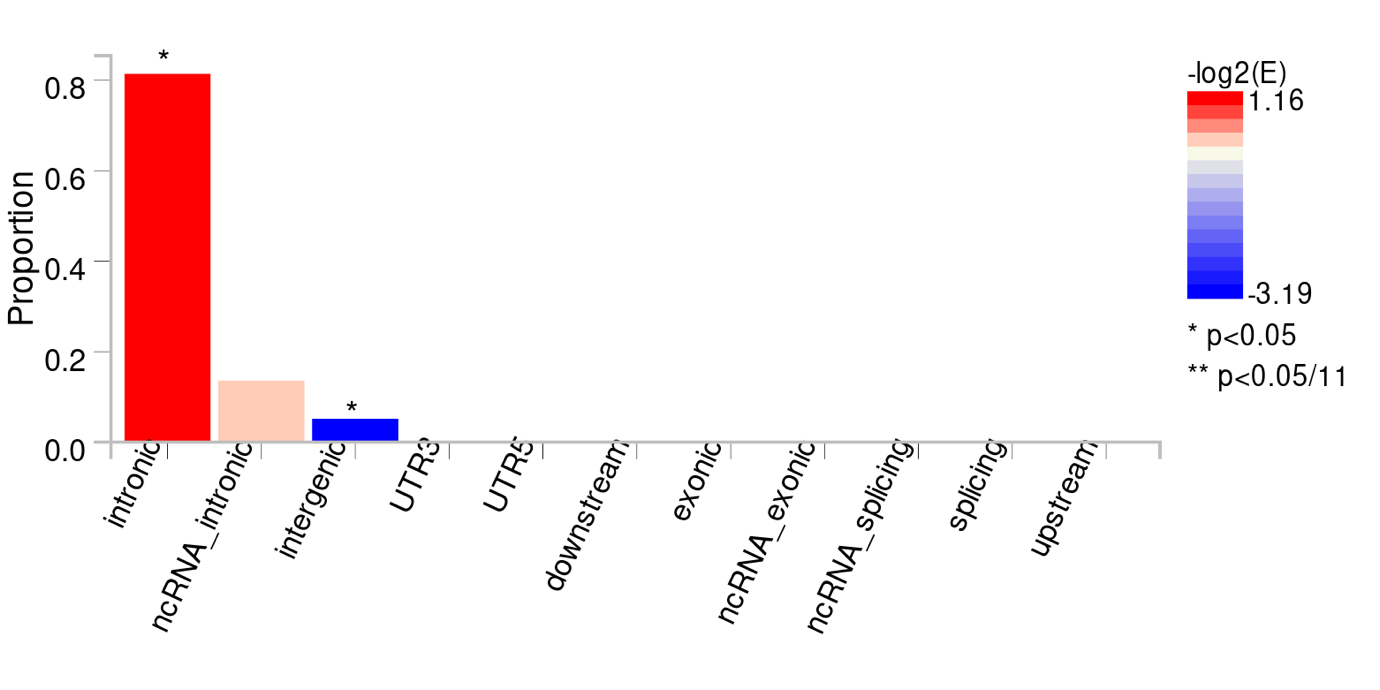

**Fig S8.** Functional consequences of 59 SNPs on genes. The plot shows the proportion of SNPs located in various genomic regions. The color gradient represents the -log2 enrichment (E) of SNPs in each category, with red indicating enrichment and blue indicating depletion. Asterisks denote statistically significant categories (*p < 0.05, **p < 0.05/11).

A)
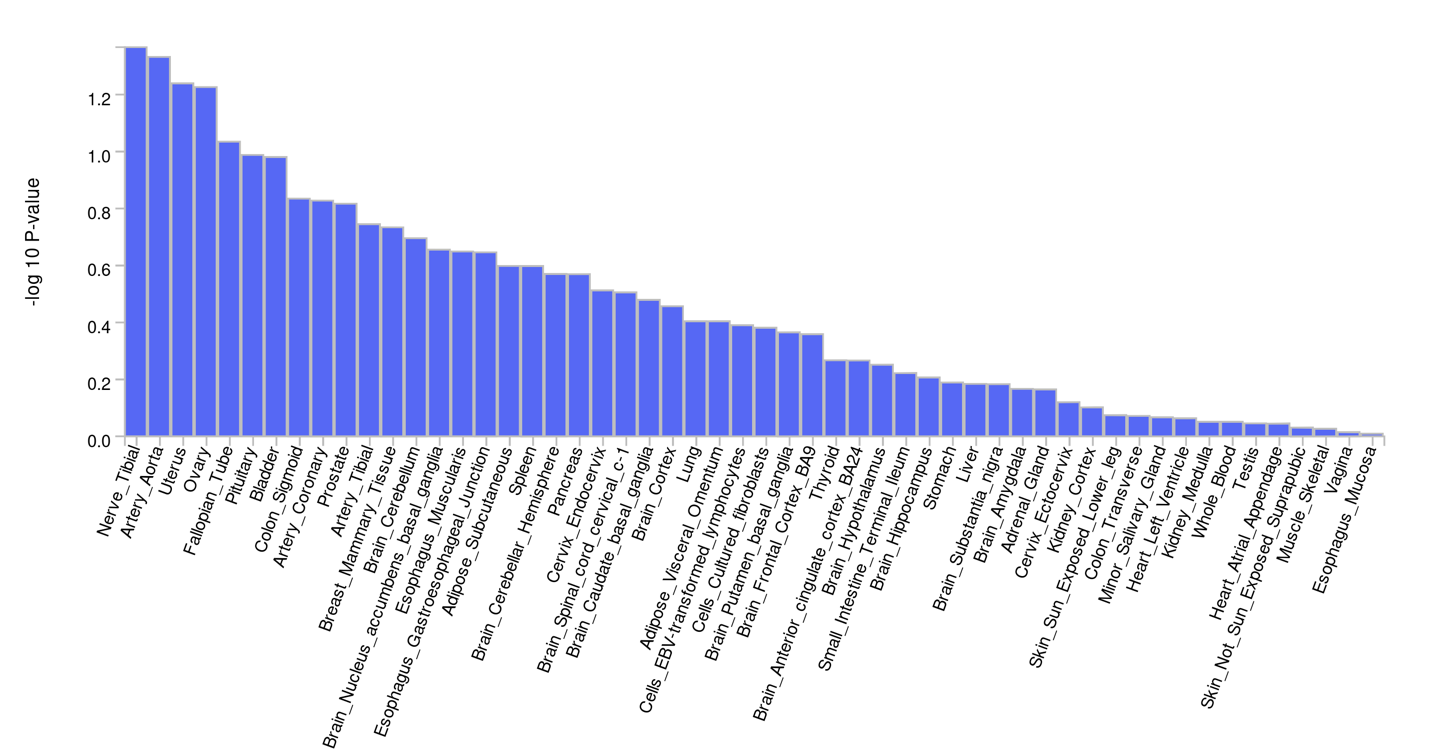

B)

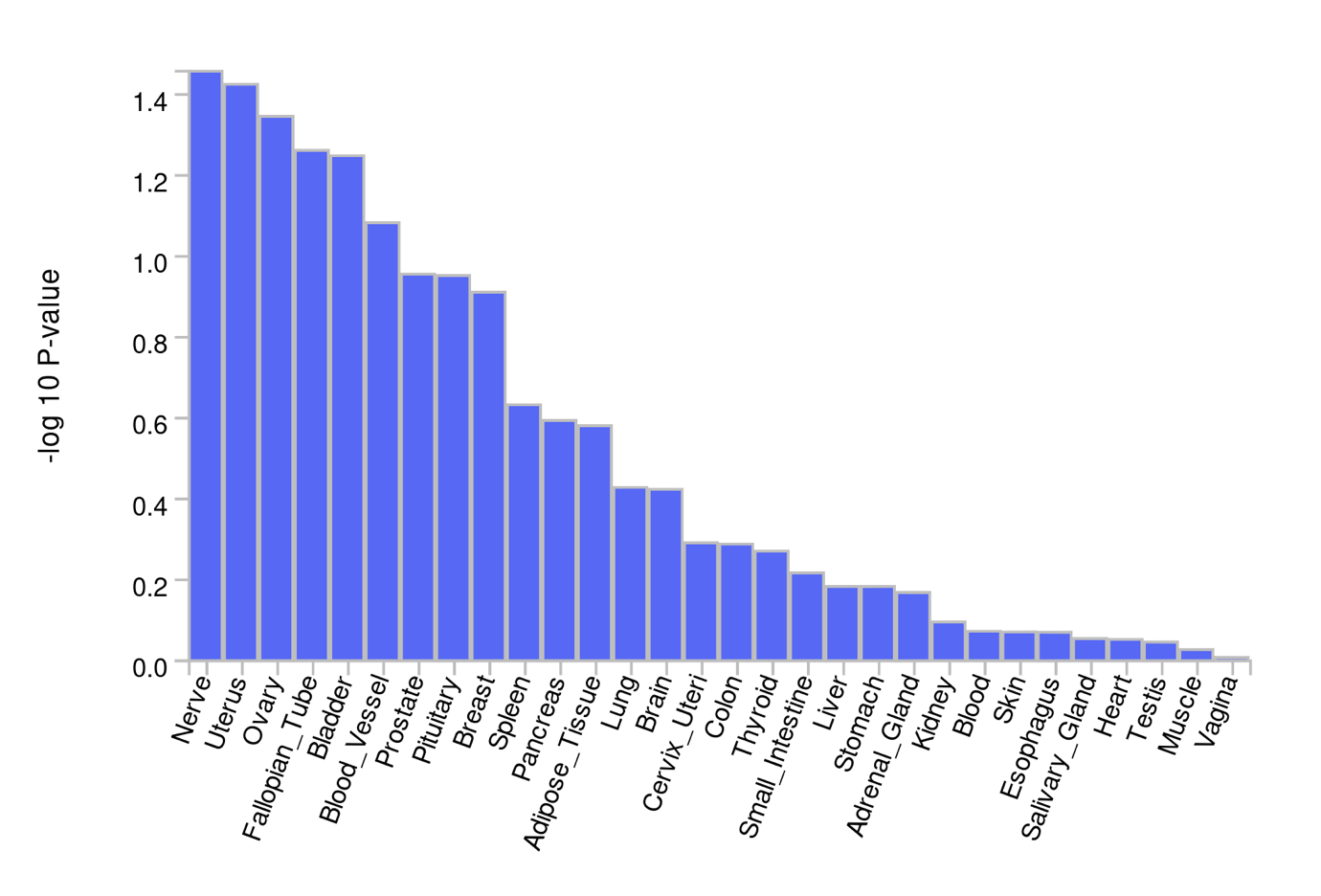

**Figure S9.** MAGMA Tissue Expression Analysis. Panel A displays the association of 53 specific tissue types with the analyzed trait based on -log10(p-values), indicating their relative enrichment. Panel B summarizes these findings across 30 general tissue types, highlighting broader tissue categories with significant associations. The x-axis represents tissue types, while the y-axis shows statistical significance (-log10 p-value) for their involvement.

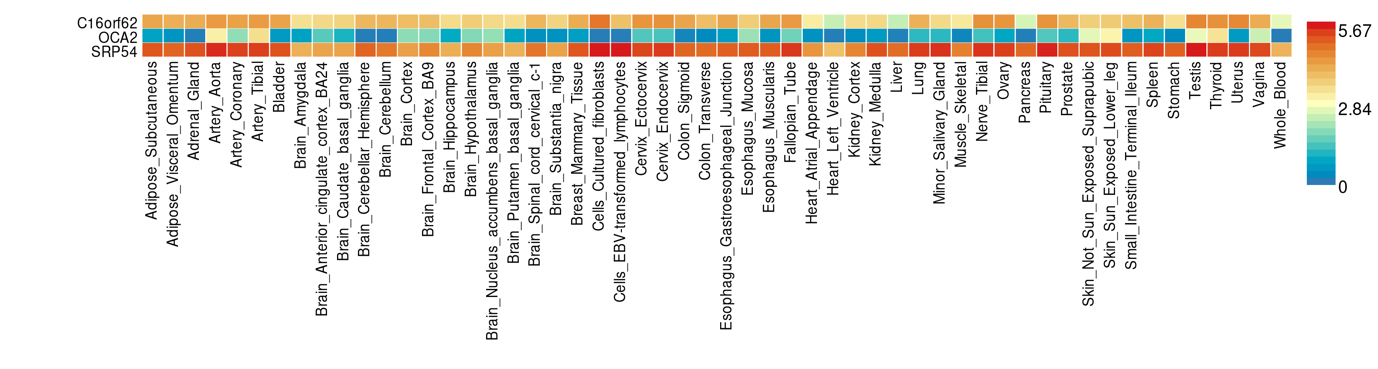

**Fig. S10.** Heat map of tissue‐specific gene expression of genes corresponding to genetic variants from meta‐analysis of pre-term and term ASD individuals. The heat map is plotted using FUMA v1.5.0 and gene expression data from GTEx v8 54 tissue types. The heat map was ordered by both gene and tissue clustering. Darker red represent higher expression of that gene compared to darker blue color across genes and tissues.

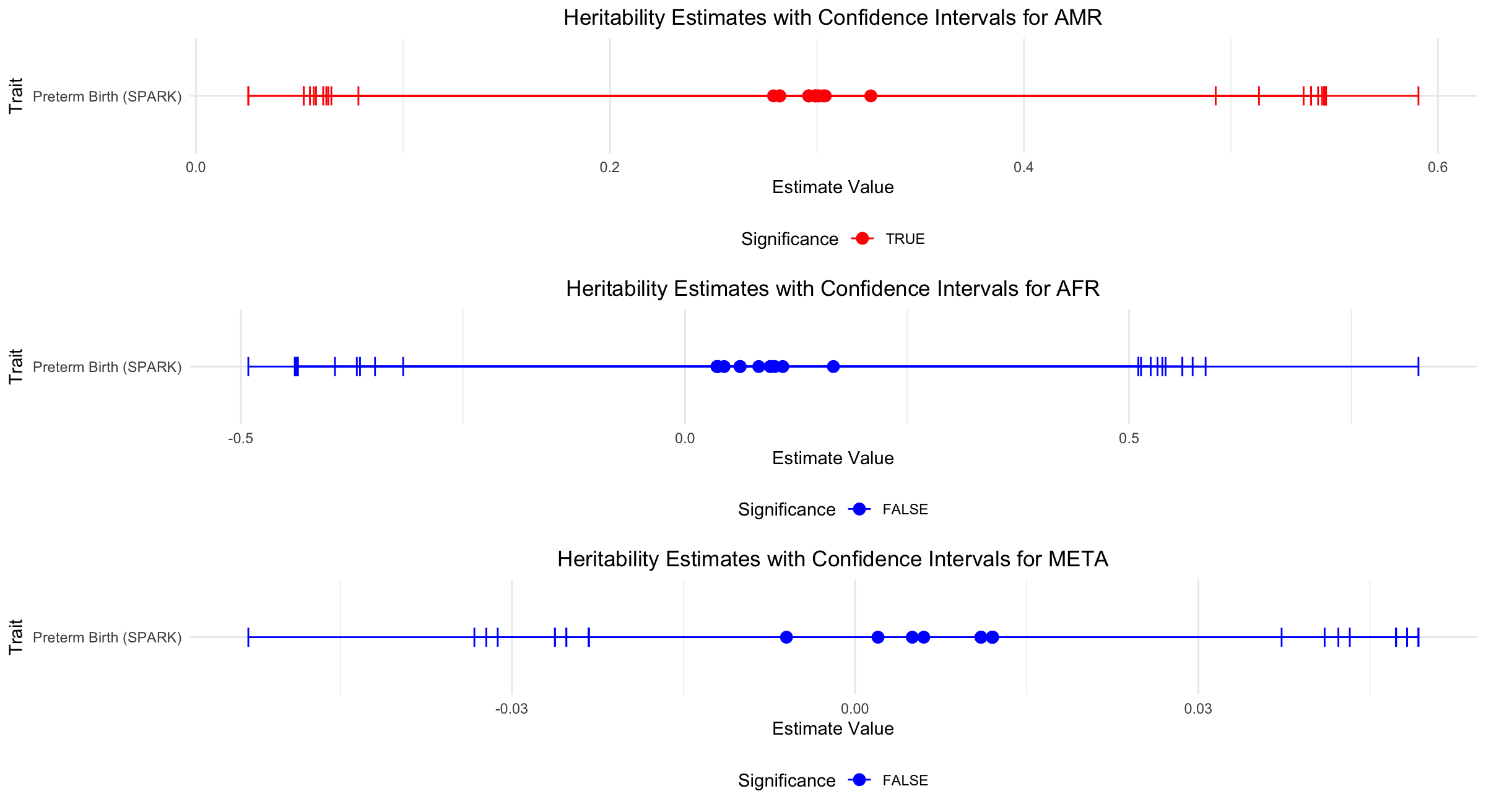

**Figure S11. Heritability estimates for Preterm birth in the individuals of AMR (A), AFR (B) ancestry and the meta-analysis (C).** Significance is indicated by red dots, while blue dots represent non-significant results with its corresponding confidence intervals. Preterm birth (SPARK) has multiple heritability estimates because its heritability was calculated multiple times using different models, subsets of individuals, or through repeat runs. This variability can occur when analyzing the same data with slightly different assumptions or subsets. However, even though these estimates are different, they are close to each other, indicating consistency in the heritability estimates despite the use of varying methods or data subsets.

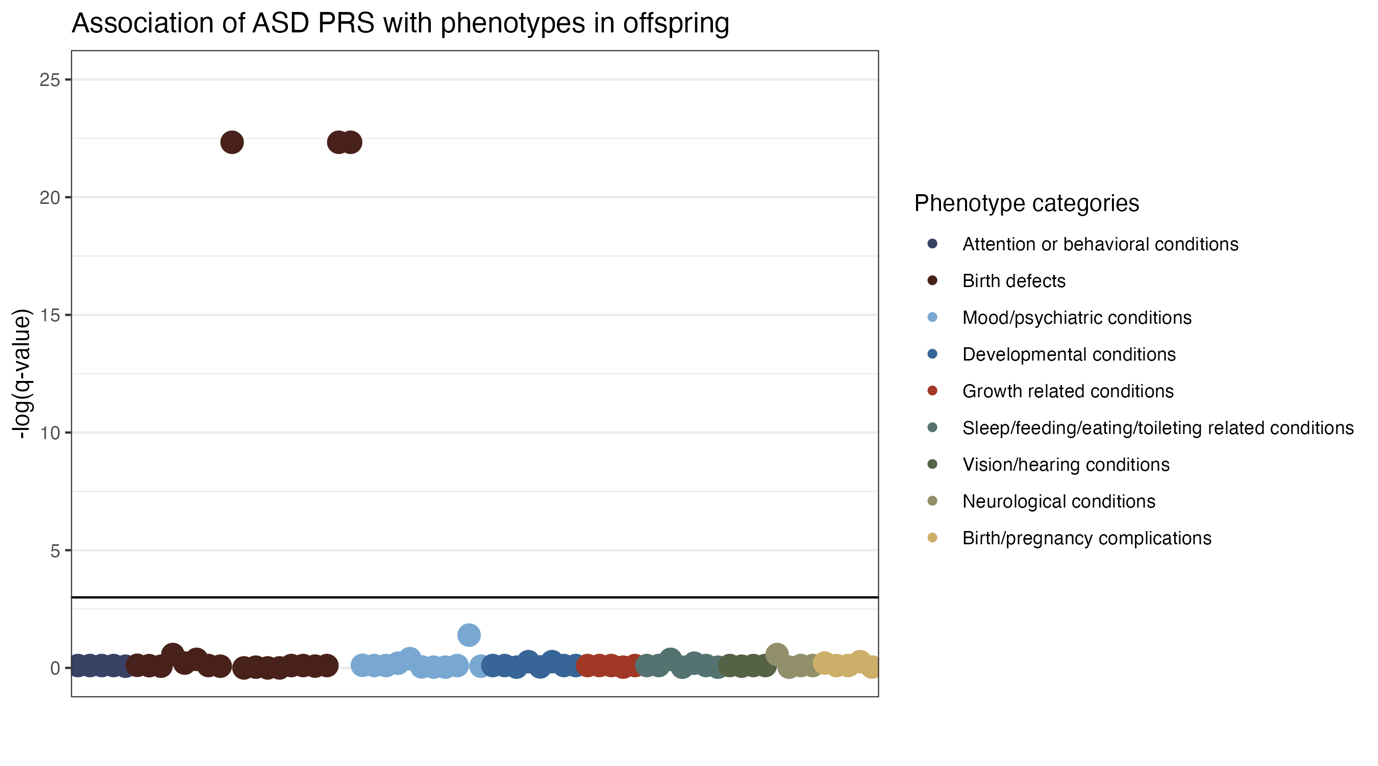

**Figure S12.** Associations between polygenic liability to preterm birth (PRS) and various medical conditions (see Table S5 for regression parameters).

**A)**
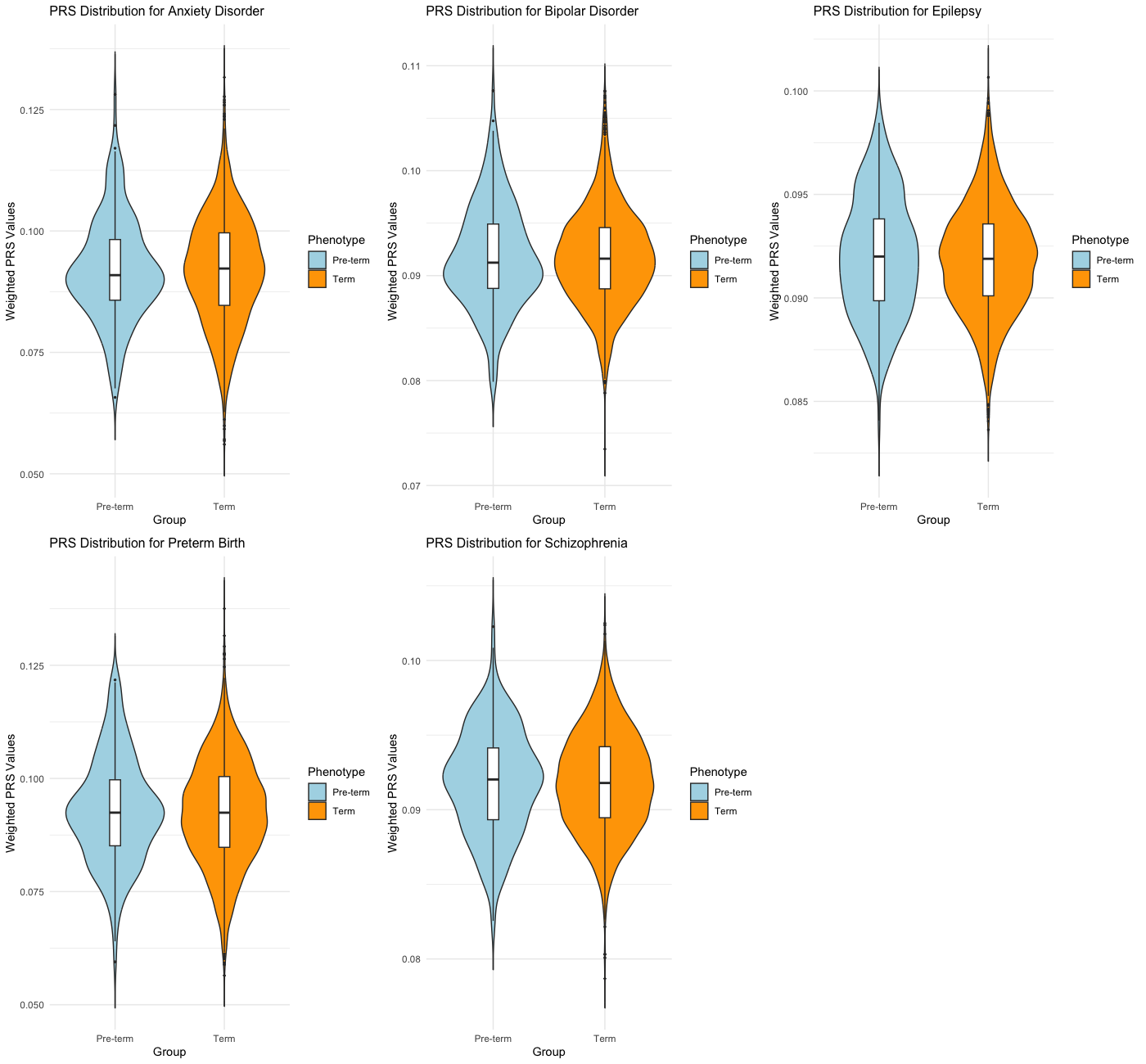

**B)**
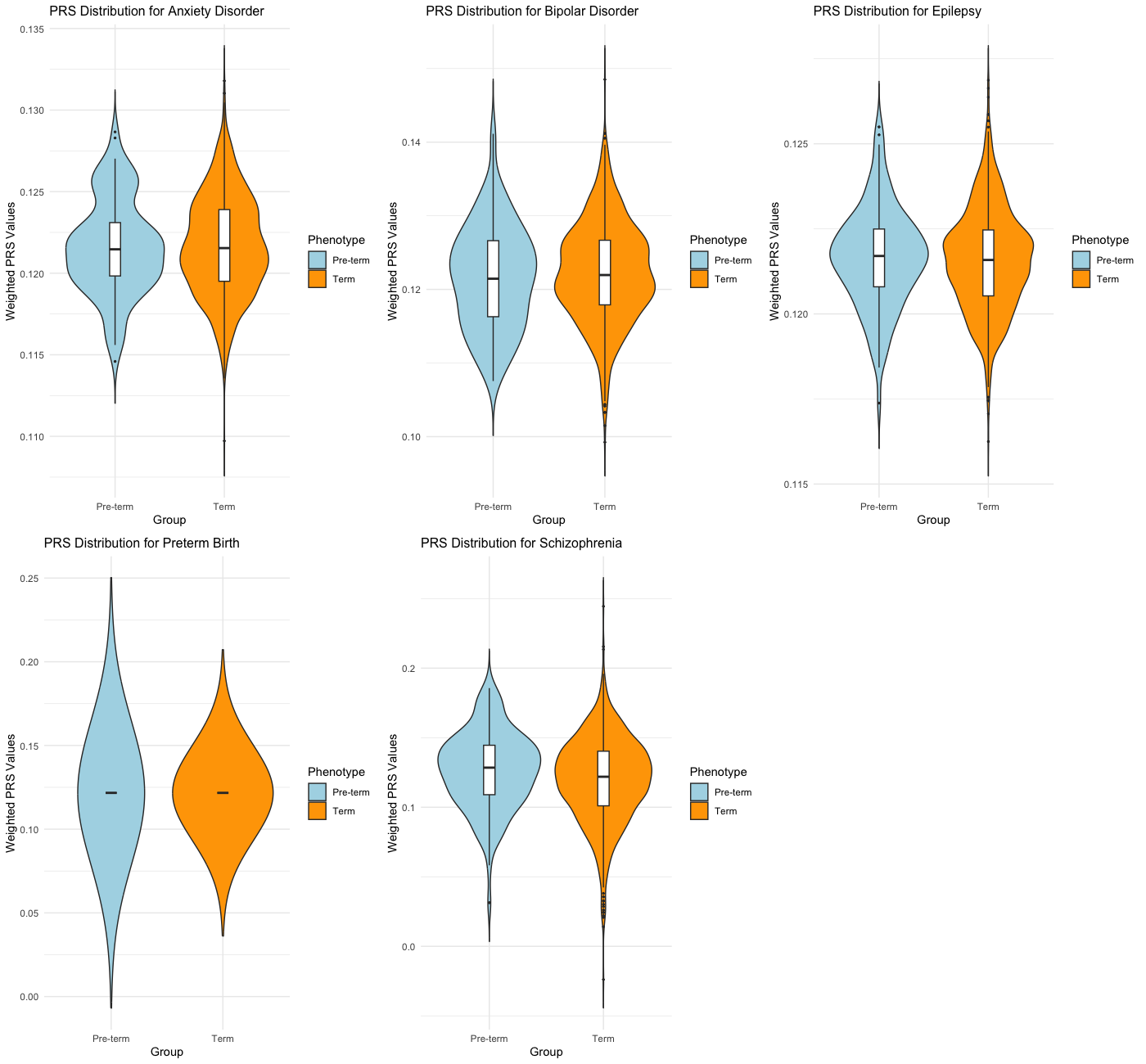

**C)**
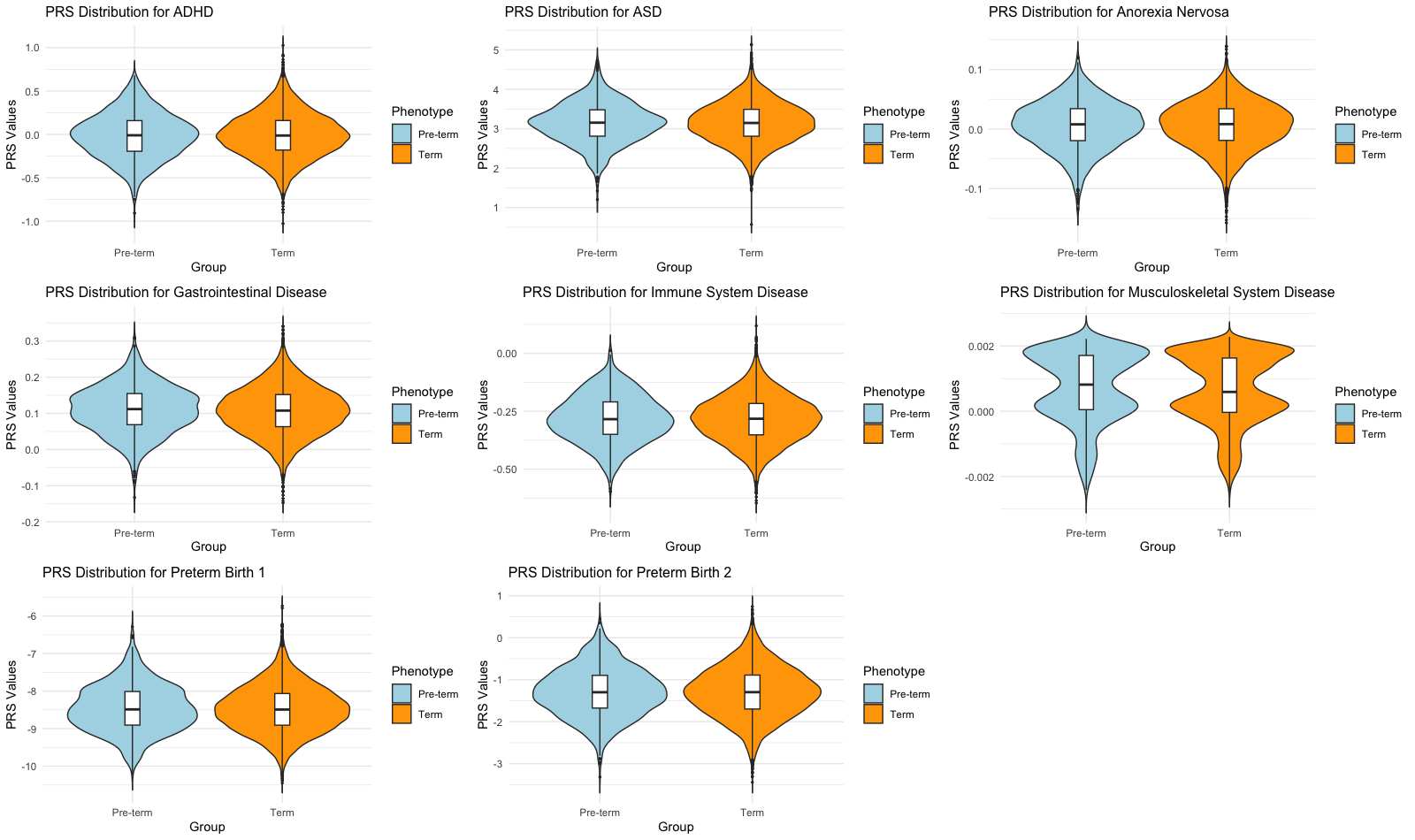

**Figure S13.** Violin plots displaying the distribution of PRS for different phenotypes in preterm (blue) and term (orange) groups of AMR (A), AFR (B) and EUR (C) ancestry. The distribution of PRS values is depicted using violin plots, with the width indicating the density of scores at different levels. Boxplots overlaid on each violin plot provide a summary of the PRS distribution, showing the median, quartiles, and outliers.

**Supplemental Tables**

| **Trait** | **AFR** | **AMR** | **EUR** |
| --- | --- | --- | --- |
| **Preterm Birth** | AFR (Rappoport et al., 2018) https://doi.org/10.1038/s41598-017-18246-5 | [AMR (Rappoport et al., 2018) https://doi.org/10.1038/s41598-017-18246-5](https://doi.org/10.1038/s41598-017-18246-5) | EUR 1 (Liu et al., 2019) doi:10.1038/s41467-019-11881-8 & EUR 2 (Rappoport et al., 2018) https://doi.org/10.1038/s41598-017-18246-5 |
| **Bipolar disorder** | AFR (O’Connell et al., 2024) https://doi.org/10.1101/2023.10.07.23296687 | Trans-ancestry (O’Connell et al., 2024) https://doi.org/10.1101/2023.10.07.23296687 | [EUR (Mullins et al., 2021) DOI: 10.1038/s41588-021-00857-4](https://doi.org/10.1038/s41588-021-00857-4) |
| **Anxiety Disorders** | AFR (Friligkou et al., 2024) doi: 10.1038/s41588-024-01908-2 | AMR (Friligkou et al., 2024) doi: 10.1038/s41588-024-01908-2 | EUR (Friligkou et al., 2024) doi: 10.1038/s41588-024-01908-2 |
| **SCZ** | AFR (Trubetskoy et al., 2022) https://doi.org/10.1038/s41586-022-04434-5 | [Trans-ancestry (Trubetskoy et al., 2022) https://doi.org/10.1038/s41586-022-04434-5](https://figshare.com/ndownloader/files/34517861) | FinnGen (Kurki et al., 2023) |
| **Epilepsy** | [Trans-ancestry (ILAE, 2023) https://doi.org/10.1038/s41588-023-01485-w](https://doi.org/10.1038/s41588-023-01485-w) | [Trans-ancestry (ILAE, 2023) https://doi.org/10.1038/s41588-023-01485-w](https://doi.org/10.1038/s41588-023-01485-w) | Trans-ancestry (ILAE, 2014) DOI: 10.1016/S1474-4422(14)70171-1 |
| **ADHD** | NA | NA | [EUR (Demontis et al., 2023) https://doi.org/10.1038/s41588-022-01285-8](https://doi.org/10.1038/s41588-022-01285-8) |
| **ASD** | NA | NA | [EUR (Grove et al., 2019) https://doi.org/10.1038/s41588-019-0344-8](https://doi.org/10.1038/s41588-019-0344-8) |
| **Anorexia Nervosa** | NA | NA | EUR (Watson et al., 2019) DOI: 10.1038/s41588-019-0439-2 |
| **Immune System Disease** | NA | NA | EUR (Donertas et al., 2021) DOI: 10.1038/s43587-021-00051-5 |
| **Musculoskeletal System Disease** | NA | NA | EUR (Donertas et al., 2021) DOI: 10.1038/s43587-021-00051-6 |
| **Gastrointestinal Disease** | NA | NA | EUR (Donertas et al., 2021) DOI: 10.1038/s43587-021-00051-7 |

**Table S1.** Dataset Details for Each Trait and Ancestry.

| **Phenotype1** | **Phenotype2** | **rg** | **se** | **z** | **p** |
| --- | --- | --- | --- | --- | --- |
| Panic Disorder | Meta | -0.789 | 0.642 | -1.229 | 0.219 |
| Gastrointestinal Disease | Meta | -0.053 | 0.081 | -0.660 | 0.509 |
| Panic Disorder | EUR | 0.845 | 0.788 | 1.073 | 0.284 |
| Preterm Birth (external) | Meta | -0.099 | 0.277 | -0.358 | 0.720 |
| Schizophrenia (EUR) | EUR | -0.083 | 0.036 | -2.323 | 0.020 |
| Anorexia Nervosa | Meta | -0.132 | 0.092 | -1.432 | 0.152 |
| Immune System Disease | EUR | 0.081 | 0.054 | 1.506 | 0.132 |
| Anorexia Nervosa | EUR | 0.080 | 0.071 | 1.117 | 0.264 |
| Immune System Disease | Meta | -0.066 | 0.086 | -0.770 | 0.441 |
| ASD | EUR | -0.117 | 0.072 | -1.615 | 0.106 |
| Musculoskeletal System Disease | EUR | 0.070 | 0.049 | 1.441 | 0.150 |
| Bipolar Disorder | EUR | -0.082 | 0.044 | -1.889 | 0.059 |
| Gastrointestinal Disease | EUR | 0.004 | 0.052 | 0.073 | 0.942 |
| Schizophrenia (all) | EUR | -0.062 | 0.031 | -1.995 | 0.046 |
| Musculoskeletal System Disease | Meta | -0.052 | 0.074 | -0.701 | 0.483 |
| Preterm Birth (external) | EUR | 0.105 | 0.174 | 0.602 | 0.548 |
| Schizophrenia (all) | Meta | 0.090 | 0.050 | 1.805 | 0.071 |
| Bipolar Disorder | Meta | 0.117 | 0.071 | 1.650 | 0.099 |
| ADHD | Meta | 0.057 | 0.070 | 0.808 | 0.419 |
| Epilepsy | EUR | 0.009 | 0.086 | 0.106 | 0.916 |
| ASD | Meta | 0.204 | 0.115 | 1.770 | 0.077 |
| Schizophrenia (EUR) | Meta | 0.135 | 0.053 | 2.537 | 0.011 |
| ADHD | EUR | 0.025 | 0.043 | 0.584 | 0.559 |
| Epilepsy | Meta | 0.031 | 0.124 | 0.251 | 0.802 |

**Table S2.** LDSC analysis results in the full sample (preterm full) and restricted to individuals of European (EUR) ancestry.

| **Phenotype** | **Model** | **Val_obs** | **SE** | **Z** | **P(Z)** |
| --- | --- | --- | --- | --- | --- |
| Preterm Birth (SPARK) | h1^2 | 0.296 | 0.122 | 2.428 | 0.015 |
| ADHD | h2^2 | 0.044 | 0.005 | 8.064 | 0.000 |
| ADHD | pge | -0.068 | 0.162 | 6.595 | 0.000 |
| Preterm Birth (SPARK) | h1^2 | 0.282 | 0.131 | 2.154 | 0.031 |
| Panic Disorder | h2^2 | 0.032 | 0.030 | 1.076 | 0.282 |
| Panic Disorder | pge | 0.556 | 0.643 | 0.691 | 0.490 |
| Preterm Birth (SPARK) | h1^2 | 0.279 | 0.109 | 2.560 | 0.010 |
| ASD | h2^2 | 0.140 | 0.039 | 3.625 | 0.000 |
| ASD | pge | -0.146 | 0.232 | 4.935 | 0.000 |
| Preterm Birth (SPARK) | h1^2 | 0.300 | 0.125 | 2.405 | 0.016 |
| Bipolar Disorder | h2^2 | 0.045 | 0.005 | 8.557 | 0.000 |
| Bipolar Disorder | pge | -0.130 | 0.150 | 7.546 | 0.000 |
| Preterm Birth (SPARK) | h1^2 | 0.303 | 0.122 | 2.482 | 0.013 |
| Anorexia Nervosa | h2^2 | 0.105 | 0.016 | 6.579 | 0.000 |
| Anorexia Nervosa | pge | 0.066 | 0.168 | 5.561 | 0.000 |
| Preterm Birth (SPARK) | h1^2 | 0.326 | 0.135 | 2.421 | 0.015 |
| Epilepsy | h2^2 | 0.080 | 0.014 | 5.567 | 0.000 |
| Epilepsy | pge | 0.190 | 0.228 | 3.552 | 0.000 |
| Preterm Birth (SPARK) | h1^2 | 0.304 | 0.123 | 2.476 | 0.013 |
| Gastrointestinal Disease | h2^2 | 0.013 | 0.002 | 5.231 | 0.000 |
| Gastrointestinal Disease | pge | 0.162 | 0.236 | 3.555 | 0.000 |
| Preterm Birth (SPARK) | h1^2 | 0.296 | 0.111 | 2.675 | 0.007 |
| Immune System Disease | h2^2 | 0.027 | 0.008 | 3.244 | 0.001 |
| Immune System Disease | pge | 0.069 | 0.136 | 6.866 | 0.000 |
| Preterm Birth (SPARK) | h1^2 | 0.282 | 0.131 | 2.154 | 0.031 |
| Musculoskeletal System Disease | h2^2 | 0.001 | 0.001 | 0.920 | 0.358 |
| Musculoskeletal System Disease | pge | 0.618 | 0.724 | 0.527 | 0.598 |
| Preterm Birth (SPARK) | h1^2 | 0.301 | 0.124 | 2.425 | 0.015 |
| Preterm Birth (external) | h2^2 | 0.008 | 0.009 | 0.819 | 0.413 |
| Preterm Birth (external) | pge | 0.760 | 0.677 | 0.355 | 0.723 |
| Preterm Birth (SPARK) | h1^2 | 0.299 | 0.126 | 2.385 | 0.017 |
| Schizophrenia (EUR) | h2^2 | 0.325 | 0.044 | 7.408 | 0.000 |
| Schizophrenia (EUR) | pge | 0.061 | 0.127 | 7.373 | 0.000 |
| Preterm Birth (SPARK) | h1^2 | 0.299 | 0.126 | 2.384 | 0.017 |
| Schizophrenia (all) | h2^2 | 0.245 | 0.024 | 10.006 | 0.000 |
| Schizophrenia (all) | pge | 0.055 | 0.127 | 7.465 | 0.000 |

**Table S3.** Popcorn results of genetic correlations between preterm birth and additional phenotypes in individuals of Admixed American (AMR) ancestry.

| **Phenotype** | **Model** | **Val_obs** | **SE** | **Z** | **P(Z)** |
| --- | --- | --- | --- | --- | --- |
| Preterm Birth (SPARK) | h1^2 | 0.096 | 0.250 | 0.384 | 0.701 |
| ADHD | h2^2 | 0.054 | 0.010 | 5.247 | 0.000 |
| ADHD | pge | -0.579 | 0.947 | 1.667 | 0.095 |
| Preterm Birth (SPARK) | h1^2 | 0.062 | 0.254 | 0.243 | 0.808 |
| Panic Disorder | h2^2 | 0.088 | 0.056 | 1.555 | 0.120 |
| Panic Disorder | pge | -1.798 | 5.158 | 0.543 | 0.587 |
| Preterm Birth (SPARK) | h1^2 | 0.110 | 0.218 | 0.503 | 0.615 |
| ASD | h2^2 | 0.120 | 0.027 | 4.463 | 0.000 |
| ASD | pge | -0.348 | 0.716 | 1.883 | 0.060 |
| Preterm Birth (SPARK) | h1^2 | 0.037 | 0.243 | 0.152 | 0.880 |
| Bipolar Disorder | h2^2 | 0.073 | 0.010 | 6.984 | 0.000 |
| Bipolar Disorder | pge | -0.031 | 0.721 | 1.431 | 0.152 |
| Preterm Birth (SPARK) | h1^2 | 0.096 | 0.227 | 0.422 | 0.673 |
| Anorexia Nervosa | h2^2 | 0.170 | 0.026 | 6.552 | 0.000 |
| Anorexia Nervosa | pge | 0.286 | 0.657 | 1.086 | 0.277 |
| Preterm Birth (SPARK) | h1^2 | 0.167 | 0.336 | 0.496 | 0.620 |
| Epilepsy | h2^2 | 0.083 | 0.026 | 3.213 | 0.001 |
| Epilepsy | pge | -0.316 | 0.809 | 1.627 | 0.104 |
| Preterm Birth (SPARK) | h1^2 | 0.101 | 0.240 | 0.420 | 0.674 |
| Gastrointestinal Disease | h2^2 | 0.022 | 0.005 | 4.467 | 0.000 |
| Gastrointestinal Disease | pge | -0.320 | 2.211 | 0.597 | 0.550 |
| Preterm Birth (SPARK) | h1^2 | 0.083 | 0.229 | 0.364 | 0.716 |
| Immune System Disease | h2^2 | 0.056 | 0.013 | 4.382 | 0.000 |
| Immune System Disease | pge | 0.689 | 1.217 | 0.255 | 0.799 |
| Preterm Birth (SPARK) | h1^2 | 0.062 | 0.254 | 0.243 | 0.808 |
| Musculoskeletal System Disease | h2^2 | 0.002 | 0.002 | 1.484 | 0.138 |
| Musculoskeletal System Disease | pge | -1.358 | 4.026 | 0.586 | 0.558 |
| Preterm Birth (SPARK) | h1^2 | 0.044 | 0.245 | 0.179 | 0.858 |
| Preterm Birth (external) | h2^2 | 0.011 | 0.015 | 0.720 | 0.472 |
| Preterm Birth (external) | pge | 5.252 | 28.591 | -0.149 | 0.882 |
| Preterm Birth (SPARK) | h1^2 | 0.036 | 0.242 | 0.150 | 0.881 |
| Schizophrenia (EUR) | h2^2 | 0.601 | 0.109 | 5.491 | 0.000 |
| Schizophrenia (EUR) | pge | -0.739 | 5.140 | 0.338 | 0.735 |
| Preterm Birth (SPARK) | h1^2 | 0.036 | 0.242 | 0.149 | 0.881 |
| Schizophrenia (all) | h2^2 | 0.416 | 0.061 | 6.870 | 0.000 |
| Schizophrenia (all) | pge | -0.716 | 5.290 | 0.324 | 0.746 |

**Table S4.** Popcorn results of genetic correlations between preterm birth and additional phenotypes in individuals of African/African American (AFR) ancestry.]

| **Phenotype** | **Model** | **Val_obs** | **SE** | **Z** | **P(Z)** |
| --- | --- | --- | --- | --- | --- |
| Preterm Birth (SPARK) | h1^2 | 0.002 | 0.018 | 0.129 | 0.897 |
| ADHD | h2^2 | 0.048 | 0.007 | 6.751 | 0.000 |
| ADHD | pge | 0.039 | 0.692 | 1.388 | 0.165 |
| Preterm Birth (SPARK) | h1^2 | 0.011 | 0.019 | 0.565 | 0.572 |
| Panic Disorder | h2^2 | 0.006 | 0.039 | 0.158 | 0.874 |
| Panic Disorder | pge | -4.991 | 31.495 | 0.190 | 0.849 |
| Preterm Birth (SPARK) | h1^2 | 0.005 | 0.019 | 0.266 | 0.791 |
| ASD | h2^2 | 0.178 | 0.055 | 3.217 | 0.001 |
| ASD | pge | 0.474 | 1.192 | 0.441 | 0.659 |
| Preterm Birth (SPARK) | h1^2 | 0.012 | 0.018 | 0.661 | 0.509 |
| Bipolar Disorder | h2^2 | 0.059 | 0.008 | 7.337 | 0.000 |
| Bipolar Disorder | pge | -0.167 | 0.432 | 2.702 | 0.007 |
| Preterm Birth (SPARK) | h1^2 | 0.006 | 0.019 | 0.330 | 0.742 |
| Anorexia Nervosa | h2^2 | 0.140 | 0.023 | 6.061 | 0.000 |
| Anorexia Nervosa | pge | -0.070 | 0.629 | 1.702 | 0.089 |
| Preterm Birth (SPARK) | h1^2 | -0.006 | 0.024 | -0.254 | 0.800 |
| Epilepsy | h2^2 | 0.076 | 0.019 | 4.066 | 0.000 |
| Epilepsy | pge | NaN | NaN | NaN | NaN |
| Preterm Birth (SPARK) | h1^2 | 0.012 | 0.019 | 0.646 | 0.518 |
| Gastrointestinal Disease | h2^2 | 0.020 | 0.004 | 5.414 | 0.000 |
| Gastrointestinal Disease | pge | -0.146 | 0.512 | 2.236 | 0.025 |
| Preterm Birth (SPARK) | h1^2 | 0.012 | 0.019 | 0.618 | 0.536 |
| Immune System Disease | h2^2 | 0.044 | 0.012 | 3.580 | 0.000 |
| Immune System Disease | pge | 0.160 | 0.660 | 1.273 | 0.203 |
| Preterm Birth (SPARK) | h1^2 | 0.011 | 0.019 | 0.565 | 0.572 |
| Musculoskeletal System Disease | h2^2 | 0.000 | 0.001 | 0.039 | 0.969 |
| Musculoskeletal System Disease | pge | -9.922 | 104.429 | 0.105 | 0.917 |
| Preterm Birth (SPARK) | h1^2 | 0.012 | 0.018 | 0.667 | 0.505 |
| Preterm Birth (external) | h2^2 | 0.009 | 0.012 | 0.759 | 0.448 |
| Preterm Birth (external) | pge | -1.192 | 3.635 | 0.603 | 0.546 |
| Preterm Birth (SPARK) | h1^2 | 0.012 | 0.018 | 0.661 | 0.509 |
| Schizophrenia (EUR) | h2^2 | 0.488 | 0.077 | 6.322 | 0.000 |
| Schizophrenia (EUR) | pge | -0.401 | 0.451 | 3.109 | 0.002 |
| Preterm Birth (SPARK) | h1^2 | 0.012 | 0.018 | 0.661 | 0.509 |
| Schizophrenia (all) | h2^2 | 0.348 | 0.042 | 8.338 | 0.000 |
| Schizophrenia (all) | pge | -0.226 | 0.375 | 3.270 | 0.001 |

**Table S5.** Popcorn results of heritability and genetic correlations between preterm birth and additional phenotypes in the entire sample (based on meta-analysis of ancestry-specific GWASs).

| **Category** | **Condition** | **OR [95%CI]** | **P-val** | **Q-val** | **Adj OR [95%CI]** | **Adj P-val** | **Adj Q-val** |
| --- | --- | --- | --- | --- | --- | --- | --- |
| **Attention or behavioral conditions** | ADHD (Attention Deficit-Hyperactivity Disorder) or ADD | 0.98 [0.93, 1.02] | 0.344 | 0.905 | 0.98 [0.93, 1.02] | 0.340 | 0.904 |
|  | Conduct Disorder | 1.05 [0.90, 1.21] | 0.550 | 0.905 | 1.04 [0.90, 1.21] | 0.557 | 0.912 |
|  | Intermittent Explosive Disorder | 0.96 [0.84, 1.10] | 0.574 | 0.905 | 0.95 [0.83, 1.09] | 0.472 | 0.912 |
|  | Oppositional Defiant Disorder | 1.03 [0.96, 1.12] | 0.415 | 0.905 | 1.03 [0.96, 1.12] | 0.415 | 0.912 |
|  | Tourette Syndrome or Tic Disorder | 0.98 [0.87, 1.10] | 0.732 | 0.930 | 0.97 [0.86, 1.1] | 0.670 | 0.912 |
| **Birth defects** | Clubbed foot | 1.29 [0.85, 1.96] | 0.237 | 0.896 | 1.28 [0.84, 1.96] | 0.252 | 0.904 |
|  | Missing or malformed bones | 1.08 [0.78, 1.50] | 0.641 | 0.909 | 1.08 [0.77, 1.5] | 0.658 | 0.912 |
|  | Extra fingers and/or extra toes | 1.14 [0.54, 2.39] | 0.732 | 0.930 | 1.14 [0.54, 2.4] | 0.736 | 0.944 |
|  | Spine deformity | 1.49 [1.00, 2.22] | 0.050 | 0.566 | 1.48 [0.99, 2.22] | 0.056 | 0.632 |
|  | Cleft lip | 0.82 [0.63, 1.08] | 0.159 | 0.814 | 0.82 [0.63, 1.08] | 0.159 | 0.718 |
|  | Cleft palate | 0.68 [0.44, 1.06] | 0.092 | 0.695 | 0.68 [0.44, 1.06] | 0.093 | 0.699 |
|  | Brain malformation/abnormality (shown on MRI) | 1.19 [0.85, 1.66] | 0.310 | 0.905 | 1.19 [0.85, 1.66] | 0.315 | 0.904 |
|  | Spina bifida/myelomeningocele (baby born with open spine, spinal cord outside of the body) | 1.11 [0.63, 1.95] | 0.724 | 0.930 | 1.11 [0.62, 1.96] | 0.732 | 0.944 |
|  | Esophageal atresia (no connection between the esophagus and stomach) | 0.69 [0.67, 0.70] | 0.000 | 0.000 | 0.67 [0.64, 0.7] | 0.000 | 0.000 |
|  | Hirschsprung disease | 1.00 [0.98, 1.02] | 1.000 | 1.000 | 1.00 [0.98, 1.02] | 1.000 | 1.000 |
|  | Intestinal malrotation | 0.94 [0.53, 1.67] | 0.839 | 0.969 | 0.94 [0.54, 1.62] | 0.810 | 0.950 |
|  | Pyloric stenosis (blockage from stomach to small intestine) | 1.00 [0.98, 1.02] | 1.000 | 1.000 | 1.00 [0.98, 1.02] | 1.000 | 1.000 |
|  | Congenital diaphragmatic hernia | 1.00 [0.98, 1.02] | 1.000 | 1.000 | 1.00 [0.98, 1.02] | 1.000 | 1.000 |
|  | Congenital heart disease/defect | 0.94 [0.75, 1.19] | 0.620 | 0.905 | 0.94 [0.75, 1.19] | 0.617 | 0.912 |
|  | Lung malformation | 0.93 [0.75, 1.17] | 0.545 | 0.905 | 0.93 [0.73, 1.18] | 0.544 | 0.912 |
|  | Hypospadias (in boys, urinary opening is in the wrong place) | 1.08 [0.76, 1.55] | 0.668 | 0.926 | 1.08 [0.76, 1.55] | 0.659 | 0.912 |
|  | Kidney malformation (for example horseshoe kidney) | 0.88 [0.53, 1.46] | 0.614 | 0.905 | 0.87 [0.52, 1.45] | 0.593 | 0.912 |
|  | Missing kidney | 2.36 [2.24, 2.48] | 0.000 | 0.000 | 2.38 [2.25, 2.52] | 0.000 | 0.000 |
|  | Missing uterus | 0.41 [0.39, 0.42] | 0.000 | 0.000 | 0.42 [0.34, 0.51] | 0.000 | 0.000 |
| **Birth/pregnancy complications** | Fetal Alcohol Syndrome, alcohol or drug exposure in mother's pregnancy | 1.15 [0.94, 1.40] | 0.168 | 0.814 | 1.15 [0.94, 1.4] | 0.169 | 0.718 |
|  | Bleed into the brain | 1.12 [0.87, 1.44] | 0.393 | 0.905 | 1.12 [0.87, 1.44] | 0.400 | 0.912 |
|  | Insufficient oxygen at birth with NICU stay | 0.96 [0.86, 1.06] | 0.421 | 0.905 | 0.96 [0.86, 1.06] | 0.421 | 0.912 |
|  | Serious prenatal infection (for example, German measles) | 1.34 [0.92, 1.95] | 0.123 | 0.767 | 1.34 [0.92, 1.94] | 0.123 | 0.718 |
|  | Premature birth (delivery before 37 weeks) | 1.01 [0.94, 1.09] | 0.808 | 0.963 | 1.01 [0.94, 1.09] | 0.805 | 0.950 |
| **Developmental conditions** | Cognitive delays or impairment due to another medical condition or exposure (For example, brain injury, stroke, lead poisoning, FAS, HIV, radiation, hydrocephalus, brain tumor, drug effects, etc.) | 1.03 [0.93, 1.13] | 0.609 | 0.905 | 1.02 [0.93, 1.13] | 0.629 | 0.912 |
|  | Intellectual disability, cognitive impairment, global developmental delay, or borderline intellectual functioning | 1.01 [0.96, 1.07] | 0.616 | 0.905 | 1.01 [0.96, 1.07] | 0.607 | 0.912 |
|  | Language delay or language disorder | 1.00 [0.95, 1.04] | 0.872 | 0.969 | 1.00 [0.95, 1.05] | 0.883 | 0.969 |
|  | Learning disability (LD, learning disorder, including reading, written expression, math, or NVLD (Nonverbal learning disability)) | 0.96 [0.91, 1.01] | 0.129 | 0.767 | 0.96 [0.91, 1.01] | 0.123 | 0.718 |
|  | Motor delay (e.g., delay in walking) or developmental coordination disorder | 0.99 [0.94, 1.05] | 0.784 | 0.959 | 0.99 [0.94, 1.05] | 0.783 | 0.950 |
|  | Mutism | 1.18 [0.95, 1.47] | 0.135 | 0.767 | 1.18 [0.95, 1.47] | 0.140 | 0.718 |
|  | Social (Pragmatic) Communication Disorder | 1.03 [0.97, 1.09] | 0.393 | 0.905 | 1.03 [0.97, 1.1] | 0.346 | 0.904 |
|  | Speech articulation problems | 0.98 [0.93, 1.04] | 0.554 | 0.905 | 0.98 [0.93, 1.04] | 0.543 | 0.912 |
|  | Cognitive delays or impairment due to another medical condition or exposure (For example, brain injury, stroke, lead poisoning, FAS, HIV, radiation, hydrocephalus, brain tumor, drug effects, etc.) | 1.03 [0.93, 1.13] | 0.609 | 0.905 | 1.02 [0.93, 1.13] | 0.629 | 0.912 |
| **Growth related conditions** | Difficulty gaining weight | 1.04 [0.92, 1.18] | 0.494 | 0.905 | 1.04 [0.92, 1.18] | 0.496 | 0.912 |
|  | Large head size (macrocephaly) | 1.05 [0.93, 1.20] | 0.416 | 0.905 | 1.06 [0.94, 1.21] | 0.339 | 0.904 |
|  | Small head size (microcephaly) | 1.11 [0.92, 1.34] | 0.284 | 0.905 | 1.11 [0.92, 1.33] | 0.291 | 0.904 |
|  | Obesity | 0.99 [0.87, 1.12] | 0.889 | 0.969 | 0.98 [0.86, 1.11] | 0.704 | 0.938 |
|  | Short stature | 1.08 [0.94, 1.25] | 0.286 | 0.905 | 1.08 [0.94, 1.25] | 0.288 | 0.904 |
| **Mood/psychiatric conditions** | Alcohol or Substance Use | 0.87 [0.68, 1.10] | 0.236 | 0.896 | 0.81 [0.62, 1.07] | 0.138 | 0.718 |
|  | Anxiety disorder, such as panic, phobia, agoraphobia, or generalized anxiety disorder (GAD) except for social anxiety | 0.99 [0.94, 1.04] | 0.625 | 0.905 | 0.98 [0.93, 1.04] | 0.480 | 0.912 |
|  | Bipolar (Manic-Depressive) Disorder | 1.03 [0.92, 1.16] | 0.574 | 0.905 | 1.02 [0.90, 1.15] | 0.768 | 0.950 |
|  | Depression or dysthymia | 1.04 [0.98, 1.11] | 0.192 | 0.818 | 1.04 [0.97, 1.12] | 0.307 | 0.904 |
|  | Disruptive Mood Dysregulation Disorder | 1.12 [0.99, 1.27] | 0.069 | 0.675 | 1.12 [0.99, 1.27] | 0.068 | 0.662 |
|  | Hoarding | 1.02 [0.89, 1.17] | 0.790 | 0.959 | 1.01 [0.88, 1.16] | 0.846 | 0.959 |
|  | Obsessive-Compulsive Disorder | 1.00 [0.94, 1.07] | 0.902 | 0.969 | 1.00 [0.93, 1.07] | 0.968 | 1.000 |
|  | Separation Anxiety | 0.99 [0.91, 1.08] | 0.871 | 0.969 | 0.99 [0.91, 1.08] | 0.850 | 0.959 |
|  | Social Anxiety Disorder/Social Phobia | 0.98 [0.92, 1.04] | 0.439 | 0.905 | 0.97 [0.90, 1.03] | 0.319 | 0.904 |
|  | Personality Disorder | 1.28 [1.05, 1.56] | 0.015 | 0.248 | 1.28 [1.02, 1.61] | 0.032 | 0.552 |
|  | Schizophrenia, Other Psychosis or Schizoaffective Disorder | 1.04 [0.86, 1.25] | 0.713 | 0.930 | 1.02 [0.84, 1.24] | 0.808 | 0.950 |
| **Neurological conditions** | Brain infection such as bacterial meningitis, encephalitis | 0.72 [0.52, 1.00] | 0.050 | 0.566 | 0.71 [0.51, 0.99] | 0.046 | 0.622 |
|  | Lead poisoning | 1.02 [0.70, 1.49] | 0.907 | 0.969 | 1.01 [0.69, 1.48] | 0.940 | 0.999 |
|  | Seizure disorder or epilepsy | 0.97 [0.89, 1.06] | 0.524 | 0.905 | 0.97 [0.89, 1.06] | 0.508 | 0.912 |
|  | Traumatic brain injury (hospitalized) | 0.89 [0.69, 1.15] | 0.377 | 0.905 | 0.88 [0.68, 1.14] | 0.343 | 0.904 |
| **Sleep/feeding/eating/**  **toileting related conditions** | Problems with eating foods - not diagnosed by a professional | 1.01 [0.96, 1.06] | 0.594 | 0.905 | 1.01 [0.96, 1.06] | 0.664 | 0.912 |
|  | Eating Disorder | 0.91 [0.70, 1.17] | 0.450 | 0.905 | 0.88 [0.66, 1.18] | 0.397 | 0.912 |
|  | Encopresis (has bowel accidents beyond age expected) | 1.08 [0.99, 1.17] | 0.084 | 0.695 | 1.08 [0.99, 1.17] | 0.083 | 0.699 |
|  | Enuresis (wets self beyond age expected) | 1.00 [0.93, 1.09] | 0.928 | 0.971 | 1.00 [0.92, 1.09] | 0.927 | 0.999 |
|  | Feeding/eating problems | 0.96 [0.90, 1.02] | 0.193 | 0.818 | 0.95 [0.89, 1.02] | 0.163 | 0.718 |
|  | Sleep Disorder or sleep problem diagnosed by a professional | 1.02 [0.96, 1.07] | 0.535 | 0.905 | 1.02 [0.96, 1.07] | 0.579 | 0.912 |
|  | Sleep problems not diagnosed by a professional | 1.00 [0.95, 1.05] | 0.912 | 0.969 | 1.00 [0.95, 1.05] | 0.860 | 0.959 |
| **Vision/hearing conditions** | Blindness | 0.88 [0.64, 1.21] | 0.445 | 0.905 | 0.88 [0.64, 1.21] | 0.443 | 0.912 |
|  | Cataract | 0.93 [0.60, 1.43] | 0.738 | 0.930 | 0.91 [0.58, 1.42] | 0.671 | 0.912 |
|  | Deafness/hearing loss | 1.06 [0.92, 1.23] | 0.394 | 0.905 | 1.06 [0.92, 1.23] | 0.431 | 0.912 |
|  | Strabismus | 1.05 [0.93, 1.19] | 0.460 | 0.905 | 1.05 [0.93, 1.19] | 0.465 | 0.912 |

**Table S6.** Associations between preterm PRS in the SPARK data and reported medical conditions. Preterm PRS was derived using summary statistics from preterm birth GWAS in an external (population-based) sample to avoid overfitting, using BridgePRS. Adjusted estimates were additionally adjusted for child’s year of birth and sex.

| **Phenotype** | **Est** | **2.5%** | **97.5%** | **Significant** |
| --- | --- | --- | --- | --- |
| ADHD | 0.000 | 0.000 | 0.000 | FALSE |
| ASD | 0.000 | -0.001 | 0.000 | FALSE |
| Anorexia Nervosa | 0.000 | -0.001 | 0.000 | FALSE |
| Gastrointestinal Disease | 0.000 | -0.001 | 0.001 | FALSE |
| Immune System Disease | 0.000 | 0.000 | 0.000 | FALSE |
| Musculoskeletal System Disease | 0.001 | -0.001 | 0.003 | FALSE |
| Preterm Birth 1 | 0.000 | 0.000 | 0.000 | FALSE |
| Preterm Birth 2 | 0.000 | -0.001 | 0.000 | FALSE |

**Table S7.** Association between preterm status (reported) in the SPARK sample and genetic liability to additional medical conditions in individuals of European (EUR) ancestry. PRS for all traits was calculated using BridgePRS.

| **Phenotype** | **Model** | **Prob** | **Est** | **2.5** | **97.5** | **Significant** | **Ancestry** |
| --- | --- | --- | --- | --- | --- | --- | --- |
| Anxiety Disorder | Stage1 | 0.260 | -0.001 | -0.005 | 0.001 | FALSE | AFR |
| Anxiety Disorder | Stage2 | 0.358 | 0.000 | 0.000 | 0.000 | FALSE | AFR |
| Anxiety Disorder | Stage1+2 | 0.382 | 0.000 | 0.000 | 0.000 | FALSE | AFR |
| Anxiety Disorder | Weighted | 1.000 | -0.001 | -0.005 | 0.001 | FALSE | AFR |
| Bipolar Disorder | Stage1 | 0.326 | 0.000 | -0.006 | 0.004 | FALSE | AFR |
| Bipolar Disorder | Stage2 | 0.345 | 0.000 | 0.000 | 0.000 | FALSE | AFR |
| Bipolar Disorder | Stage1+2 | 0.329 | 0.000 | -0.006 | 0.004 | FALSE | AFR |
| Bipolar Disorder | Weighted | 1.000 | 0.000 | -0.006 | 0.004 | FALSE | AFR |
| Epilepsy | Stage1 | 0.246 | -0.001 | -0.005 | 0.001 | FALSE | AFR |
| Epilepsy | Stage2 | 0.538 | 0.000 | 0.000 | 0.000 | FALSE | AFR |
| Epilepsy | Stage1+2 | 0.216 | 0.000 | 0.000 | 0.000 | FALSE | AFR |
| Epilepsy | Weighted | 1.000 | -0.001 | -0.005 | 0.001 | FALSE | AFR |
| Preterm Birth (external) | Stage1 | 0.301 | 0.000 | 0.000 | 0.000 | FALSE | AFR |
| Preterm Birth (external) | Stage2 | 0.352 | 0.000 | 0.000 | 0.000 | FALSE | AFR |
| Preterm Birth (external) | Stage1+2 | 0.347 | 0.000 | 0.000 | 0.000 | FALSE | AFR |
| Preterm Birth (external) | Weighted | 1.000 | 0.000 | 0.000 | 0.000 | FALSE | AFR |
| Schizophrenia | Stage1 | 0.018 | -0.001 | -0.006 | 0.002 | TRUE | AFR |
| Schizophrenia | Stage2 | 0.787 | 0.004 | -0.006 | 0.012 | FALSE | AFR |
| Schizophrenia | Stage1+2 | 0.194 | 0.003 | -0.005 | 0.010 | FALSE | AFR |
| Schizophrenia | Weighted | 1.000 | 0.004 | -0.006 | 0.012 | FALSE | AFR |
| Anxiety Disorder | Stage1 | 0.210 | 0.000 | -0.002 | 0.001 | FALSE | AMR |
| Anxiety Disorder | Stage2 | 0.450 | 0.000 | -0.002 | 0.001 | FALSE | AMR |
| Anxiety Disorder | Stage1+2 | 0.339 | 0.000 | -0.002 | 0.000 | FALSE | AMR |
| Anxiety Disorder | Weighted | 1.000 | 0.000 | -0.002 | 0.000 | FALSE | AMR |
| Bipolar Disorder | Stage1 | 0.344 | 0.000 | -0.003 | 0.001 | FALSE | AMR |
| Bipolar Disorder | Stage2 | 0.423 | 0.000 | -0.002 | 0.000 | FALSE | AMR |
| Bipolar Disorder | Stage1+2 | 0.234 | 0.000 | -0.003 | 0.001 | FALSE | AMR |
| Bipolar Disorder | Weighted | 1.000 | 0.000 | -0.002 | 0.001 | FALSE | AMR |
| Epilepsy | Stage1 | 0.227 | 0.000 | -0.003 | 0.002 | FALSE | AMR |
| Epilepsy | Stage2 | 0.546 | 0.000 | -0.003 | 0.001 | FALSE | AMR |
| Epilepsy | Stage1+2 | 0.228 | 0.000 | 0.000 | 0.000 | FALSE | AMR |
| Epilepsy | Weighted | 1.000 | 0.000 | -0.003 | 0.001 | FALSE | AMR |
| Preterm Birth (external) | Stage1 | 0.223 | 0.000 | 0.000 | 0.000 | FALSE | AMR |
| Preterm Birth (external) | Stage2 | 0.502 | 0.000 | -0.002 | 0.000 | FALSE | AMR |
| Preterm Birth (external) | Stage1+2 | 0.274 | 0.000 | -0.003 | 0.002 | FALSE | AMR |
| Preterm Birth (external) | Weighted | 1.000 | 0.000 | -0.002 | 0.001 | FALSE | AMR |
| Schizophrenia | Stage1 | 0.233 | 0.000 | -0.002 | 0.001 | FALSE | AMR |
| Schizophrenia | Stage2 | 0.486 | 0.000 | -0.002 | 0.000 | FALSE | AMR |
| Schizophrenia | Stage1+2 | 0.281 | 0.000 | -0.002 | 0.000 | FALSE | AMR |
| Schizophrenia | Weighted | 1.000 | 0.000 | -0.002 | 0.000 | FALSE | AMR |

**Table S8.** Association between preterm status (reported) in the SPARK sample and genetic liability to additional medical conditions in individuals of African / African American (AFR) and Admixed American (AMR) ancestry. PRS for all traits was calculated using BridgePRS.
